# Longitudinal plasma neurofilament light chain and patient-reported outcomes as complementary markers of vincristine-associated peripheral neuropathy in adults with lymphoma: a cohort study

**DOI:** 10.64898/2026.06.28.26356741

**Authors:** Gretchen A. McNally, Grace Ji-eun Shin, Lise Worthen-Chaudhari, Patrick M. Schnell, Laura Flora, Surith Sanjay Krishna, Tim Voorhees, Robert Baiocchi, David Bond, Beth Christian, Kami Maddocks, Yazeed Sawalha, Maryam B. Lustberg

**Affiliations:** Department of Nursing, The Ohio State University, Arthur G. James Cancer Hospital, Columbus, OH, USA; The Ohio State University Comprehensive Cancer Center, Columbus, OH, USA; Department of Neurology, The Ohio State University College of Medicine, Columbus, OH, USA; Department of Dermatology, The Ohio State University College of Medicine, Columbus, OH, USA; Department of Physical Medicine and Rehabilitation, The Ohio State University College of Medicine, Columbus, OH, USA; Division of Biostatistics, College of Public Health, The Ohio State University, Columbus, OH, USA; Department of Medical Epidemiology and Biostatistics, Karolinska Institute, Stockholm, Sweden; Division of Hematology, Department of Internal Medicine, The Ohio State University College of Medicine, Columbus, OH, USA; Yale Cancer Center, Yale School of Medicine, New Haven, CT, USA

**Keywords:** Peripheral neuropathy, vincristine, patient-reported outcomes, neurofilament light chain, lymphoma, survivorship

## Abstract

Chemotherapy-induced peripheral neuropathy (CIPN) is a common neurotoxicity of cancer treatment with limited diagnostic, monitoring, and treatment options. Neurofilament light chain (NfL) is an axonal cytoskeletal protein released during neuroaxonal injury and a promising biomarker of CIPN, but prospective evidence for NfL as a marker of CIPN from vincristine-containing lymphoma chemotherapy treatment remains limited. To fill this gap, we conducted a pragmatic single-center prospective observational cohort study of adults with non-Hodgkin lymphoma (NHL) receiving first-line vincristine-containing chemotherapy to evaluate NfL dynamics across multiple pre-cycle visits and assess relationships with patient-reported and clinician-graded neuropathy measures. We followed 25 participants during 4-6 months of chemotherapy, and a small subset of those participants (n=6) for 24-42 months post-chemotherapy. Serial plasma NfL was measured and CIPN symptoms were assessed using patient- and clinician-reported measures. Longitudinal changes were analyzed using mixed-effects models. Plasma NfL increased relative to pre-cycle1 at all timepoints (all p<0.001), increasing more than threefold by pre-cycle4. Patient-reported CIPN scores and clinician-graded neuropathy also increased during treatment. Exploratory pooled visit-level analyses showed a modest NfL-CIPN association (Spearman ρ=0.393, p=0.004), while timepoint-specific, lagged, and post hoc sensitivity analyses suggested potential to predict persistent CIPN symptoms from early NfL concentrations. To our knowledge, these findings provide the first prospective evidence that NfL is sensitive to vincristine exposure in adults with NHL and may complement patient-reported symptom assessment, clinician grading, and dose-modification context in future CIPN monitoring studies.

## Introduction

Chemotherapy-induced peripheral neuropathy (CIPN) is a common and clinically consequential adverse effect of vincristine, a core chemotherapy agent within first-line lymphoma treatment regimens [1–3]. Vincristine-associated CIPN has been poorly studied across hematologic cancer populations, and the limited extant literature remains heterogeneous with respect to disease subtype, regimen, neuropathy definition, and assessment method [2–4]. This gap in knowledge regarding CIPN is particularly relevant in non-Hodgkin lymphoma (NHL), a major category of hematologic cancer that includes diverse lymphoid malignancies and is commonly treated with vincristine-containing regimens such as CHOP (cyclophosphamide, doxorubicin, vincristine, and prednisone) and infusional dose-adjusted EPOCH (etoposide, prednisone, vincristine, cyclophosphamide, and doxorubicin), both combined with or without rituximab (R)[1]. The rate at which adults with NHL develop CIPN is high, at up to 75% of patients [3–5]. Estimates vary because of differences in regimen, disease subtype, neuropathy definition, and assessment method [1–5], however, all studies in this population link neuropathy with sensory symptoms, functional limitations, impaired daily activities, reduced quality of life, and persistent survivorship burden [3, 6, 7]. The broader vincristine-induced CIPN literature describes additional manifestations including neuropathic pain, weakness, impaired proprioception, gait instability, falls, autonomic symptoms, and treatment modification [2, 3, 8, 9].

Mechanistically, vincristine interferes with microtubule assembly and axonal transport, processes essential for maintaining long peripheral axons, thereby linking vincristine exposure to the distal axonal dysfunction and injury that underlie CIPN [10–12]. During neuroaxonal injury, an axonal cytoskeletal protein called neurofilament light chain (NfL) is released and measurable in blood using highly sensitive assays [13–15]. As a blood-based marker of neuroaxonal injury, NfL offers a potentially effective biomarker of CIPN development to complement patient-reported outcomes (PRO) and clinician-graded measures that capture patient experience and symptom severity [16]. In CIPN, blood NfL has been studied most extensively in taxane-, platinum-, and antibody-drug-conjugate-associated neuropathy, with evidence that levels increase during neurotoxic treatment and may correlate with neuropathy severity or persistence in some settings [17–23]. However, evidence supporting the usefulness of NfL as a biomarker of CIPN following NHL chemotherapy is lacking.

Challenges to studying NfL as a biomarker for CIPN due to NHL treatment include its lack of specificity. Circulating concentrations can increase across a broad range of central or peripheral nervous system injuries and may also be influenced by non-neuropathy factors such as age, renal function, body size or distribution volume, and neurological comorbidities [13–15, 24–27]. In the CIPN setting, the magnitude, timing, and clinical associations of NfL changes may further vary by anticancer agent, regimen, cancer population, baseline vulnerability, cumulative exposure, and sampling schedule [15, 20, 23]. These challenges in relating NfL dynamics specifically to chemotherapy exposure support the need for longitudinal studies. Ideally, such studies would be pragmatic by aligning repeated measures with clinical visits for vincristine-containing chemotherapy infusions (hereafter, “clinically aligned visits”) in real-world treatment settings, while evaluating NfL alongside established patient-reported and clinician-assessed measures. Another challenge to validating NfL as a biomarker of CIPN is determining how clinicians should interpret a blood-based marker of neuroaxonal injury alongside the patient-reported symptoms and clinician assessments that are currently used to monitor CIPN progression. The timing of patient and clinician perceptions of symptoms can differ from biological changes in ways that are not necessarily linear. Future studies must harmonize NfL dynamics with patient- and clinician-reported outcomes to determine whether NfL provides clinically useful biological context that complements, rather than duplicates, symptom-based assessment.

Characterization of NfL dynamics among adults with NHL receiving vincristine-containing regimens is clinically important because survivorship after lymphoma treatment continues to increase, yet neuropathy symptoms persist beyond treatment completion and contribute to long-term symptom burden [3, 6], such that individuals with NHL represent a population in need of improved CIPN care. Therefore, we conducted a pragmatic single-center prospective observational cohort study of adults with NHL receiving first-line vincristine-containing CHOP or EPOCH chemotherapy. We aimed to evaluate the feasibility of clinically aligned serial plasma NfL and neuropathy outcome collection, characterize longitudinal NfL dynamics during treatment, describe concurrent patient-reported and clinician-graded neuropathy outcomes, and explore association with the European Organization for Research and Treatment of Cancer Quality of Life Questionnaire-Chemotherapy Induced Peripheral Neuropathy 20-item instrument (CIPN20), Brief Pain Inventory-Short Form (BPI-SF), National Cancer Institute Common Terminology Criteria for Adverse Events (CTCAE) neuropathy, dose modification, and longer-term patient-reported outcomes. We hypothesized that plasma NfL would increase with vincristine exposure. By characterizing NfL trajectories alongside symptom, pain, dose modification, and survivorship context, this study addresses an underexplored cancer context and provides a foundation for larger studies testing whether early NfL trajectories can inform CIPN monitoring and long-term symptom risk assessment.

## Methods

### Study Design

This was a pragmatic, single-site, prospective observational cohort study designed to assess the feasibility of the study in the target population and to conduct exploratory analyses to inform future sample size estimates. We performed the study within The Ohio State University (OSU) Arthur G. James Cancer Hospital, a tertiary NCI-designated Comprehensive Cancer Center. The protocol was approved by the OSU Institutional Review Board, and written informed consent was obtained from participants receiving first-line vincristine-containing chemotherapy, CHOP or EPOCH.

### Study Participants & Setting

Eligible participants were >18 years old, diagnosed with NHL, and initiating first-line vincristine-containing CHOP- or EPOCH-based chemotherapy. Participants were excluded if they had pre-existing neuropathy, central nervous system involvement of lymphoma, non-CIPN neurodegenerative conditions, Eastern Cooperative Oncology Group (ECOG) performance status >2, or concurrent treatment with agents known to be associated with neuropathy (e.g., brentuximab or bortezomib). Disease progression and clinical complications are anticipated in this population and were considered reasons for study discontinuation; this includes participants whose disease progression required discontinuation of the full EPOCH or CHOP protocol or transfer to hospice and/or whose ECOG status increased by >2 while on study. However, participants for whom vincristine was removed, but who otherwise continued on the CHOP or EPOCH regimen, were retained in the study. All study activities occurred within The OSU Arthur G. James Cancer Hospital or within the participant’s home environment.

### Study Timeline

Standard treatment regimen includes six chemotherapy cycles administered at 21-day intervals, generally completed within a 4–6-month period. Data collection was aligned with routine clinical care, with research assessments collected during the clinical visits, deferring to the treatment plan regarding when that visit would occur. In addition, among participants who consented to additional cross-sectional follow-up, we collected PROs only at 24-42 months (2-3.5 years), after completion of vincristine exposure for a maximum potential study duration of 4 years.

### Study Activities

#### Data collection platform

Data were collected and/or managed using Research Electronic Data Capture (REDCap) tools [28, 29] hosted by OSU.

#### Screening and enrollment

Because the study was embedded in routine lymphoma care, enrollment timing depended on clinical workflow and participant availability. Some participants were first approached after treatment initiation and therefore contributed their first research blood and PRO assessments at the next clinically aligned pre-cycle visit, most commonly before cycle 2. We accounted for this by analyzing samples based on actual collection timing rather than treating all first samples as a uniform pretreatment baseline (**Fig. 1**).

**Figure 1.**
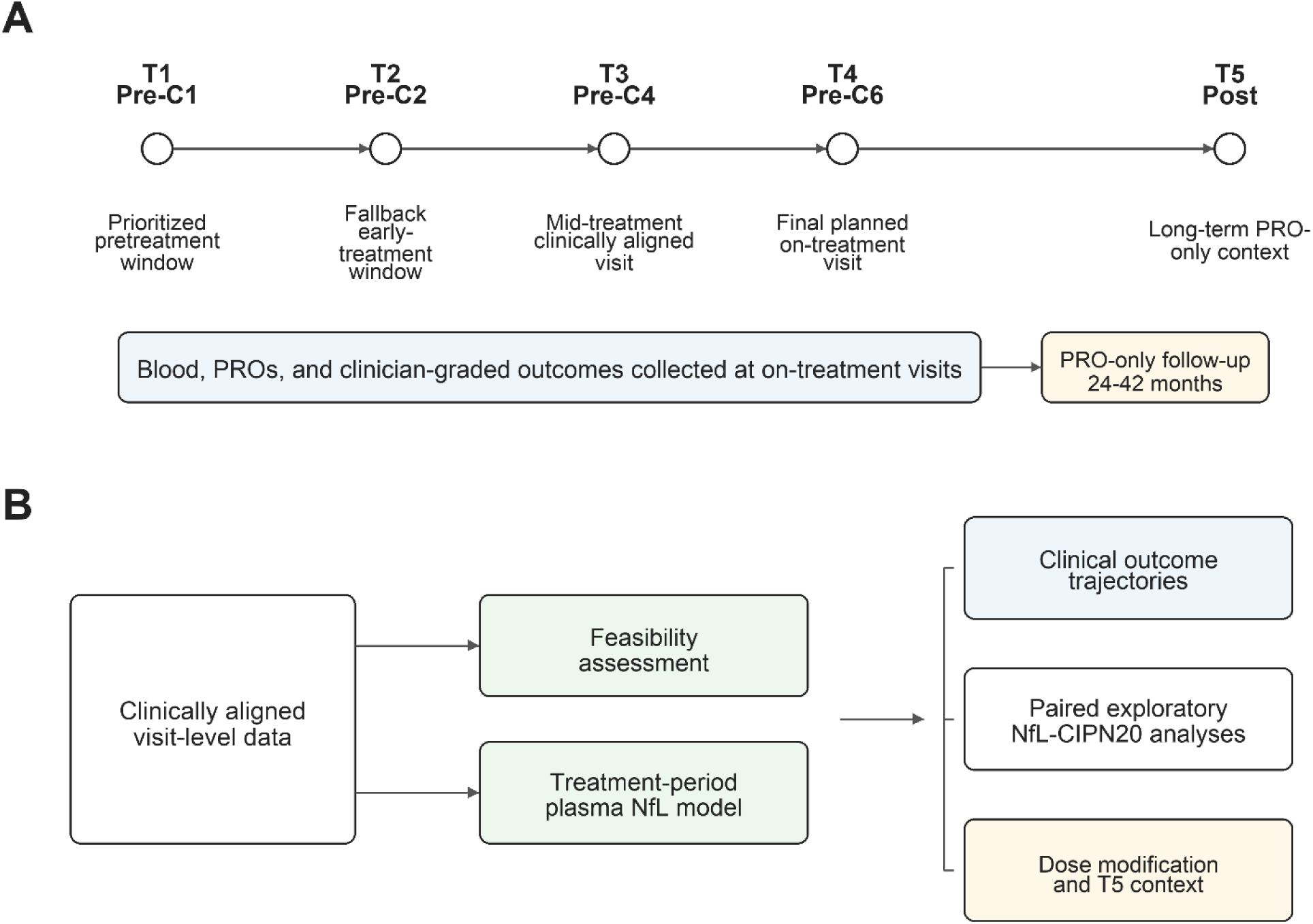
Study data collection and analysis schema. (A) Clinically aligned data-collection schema showing the five labeled timepoints used in the study: T1/Pre-C1, T2/Pre-C2, T3/Pre-C4, T4/Pre-C6, and T5/Post. T1-T4 represent planned treatment-period assessments; T5/Post represents a long-term patient-reported outcome follow-up 24-42 months after chemotherapy completion. (B) Analysis framework showing how visit-level data contributed to feasibility assessment, the treatment-period plasma NfL model, clinical outcome trajectories, paired exploratory NfL-CIPN20 analyses, and dose-modification/T5 context.

#### Medical history at enrollment and throughout study participation

Research staff extracted electronic medical record data, including height, weight, medications, comorbidities at diagnosis, clinician-reviewed outcomes, laboratory data, where available, and vincristine dose adjustments at enrollment and throughout study participation. Dose administered per cycle and dose modifications were recorded and analyzed in supplementary data. Dose modification was treated as a clinical context and a potential confounder of NfL and symptom trajectories, but was not assumed to reflect neuropathy unless a neuropathy-related reason was documented.

#### Repeated measures timepoints within routine clinical care

We collected timepoint 1 (T1) just before first chemotherapy exposure in cycle 1 (Pre-C1). As a protocol-defined fallback sampling window, we collected T2/Pre-C2 just before chemotherapy exposure in cycle 2. We prospectively prioritized T1/Pre-C1 collection while allowing T2/Pre-C2 collection as a fallback early-treatment window when T1/Pre-C1 could not be obtained. We collected T3/Pre-C4 and T4/Pre-C6 data just before the fourth and sixth cycles of chemotherapy exposure, respectively. Finally, for a subset of participants, we collected only PROs within a window of 24-42 months after completion of vincristine treatment at T5/Post. T5/Post data were analyzed descriptively and separately from the primary NfL analysis (**Fig. 1A**).

### Data Collected per Timepoint

#### Blood-based biomarker outcomes (primary)

Peripheral blood was collected from participants at each study timepoint into K3EDTA tubes. Samples were processed by the Hematology Tissue Bank at The Ohio State University within 24 hours of collection. Plasma was generated by centrifugation at ambient temperature at 2,500 rpm for 10 minutes. Plasma was aliquoted into 1-mL vials, with two 1-mL plasma aliquots created when sufficient volume was available, and stored at -80°C until needed. Frozen plasma aliquots were shipped on dry ice to Quanterix (Billerica, MA) for NfL measurement. Plasma NfL was assayed using the ultrasensitive single-molecule array Simoa NF-Light® assay (Quanterix, Billerica, MA). The limit of detection (LOD), lower limit of quantification (LLOQ), and upper limit of quantification (ULOQ) were 0.038 pg/mL, 0.174 pg/mL, and 382 pg/mL, respectively. Quality controls were included in the array and tested with participant samples. Plasma samples and controls were diluted at a 1:4 ratio and measured in duplicate with calibrators. NfL measurements were performed by Quanterix in two analytical batches.

#### Patient-reported outcomes (secondary)

The initial PRO assessment was completed with assistance from a research assistant, whereas subsequent PRO assessments were completed remotely through a REDCap survey link sent to participants by email. These measures capture patient experience of neuropathy and have demonstrated validity and responsiveness in clinical studies [30–34]. Measures collected were as follows:

1. *European Organization for Research and Treatment of Cancer Quality of Life Questionnaire-Chemotherapy Induced Peripheral Neuropathy 20-item instrument (EORTC QLQ-CIPN20)* [30–32]: The CIPN20 questionnaire was administered as the CIPN-specific PRO instrument representing sensory, motor, and autonomic neuropathy symptoms. Because the driving item and the male sexual-function item in the questionnaire were not applicable to all participants, these items were treated as not applicable when appropriate and were not imputed. We analyzed the raw applicable item CIPN20 score, calculated as the sum of completed applicable items scored from 1 (not at all) to 4 (very much). As a result, the possible minimum raw score varied across participants according to item applicability, ranging from 18 to 20, with higher scores indicating greater patient-reported symptom burden.
2. Pain severity was assessed using the *Brief Pain Inventory-Short Form (BPI-SF)* pain severity score, calculated as the mean of four items assessing pain intensity over the past 24 hours: worst pain, least pain, average pain, and current pain. Each item is rated on a 0-10 numeric rating scale, with 0 = no pain and 10 = pain as bad as the participant can imagine. The resulting severity score ranges from 0 to 10, with higher scores indicating greater pain intensity [33, 34].

#### Clinician-assessed measures (secondary)

Clinician-assessed measures were obtained from the patient chart into the REDCap database by research staff. The outcome recorded was as follows:

*1) National Cancer Institute Common Terminology Criteria for Adverse Events (NCI CTCAE v5.0):* As part of standard oncology care, treating clinicians performed routine toxicity assessments using CTCAE [35]. The study did not administer CTCAE questions directly; rather, research staff collected neuropathy-related toxicity assessment data as recorded by clinicians during routine care on a five-point scale: 0 (asymptomatic), 1 (mild), 2 (moderate), 3 (severe), 4 (life-threatening). Higher CTCAE grades indicate greater symptom severity and/or functional limitation. Because CTCAE toxicity assessment in routine practice is subject to provider-level variability, particularly for symptoms such as paresthesia that rely on clinical judgment and patient report during the visit, these data were interpreted as clinician-recorded toxicity context rather than as a standardized neurologic endpoint. Patient-reported outcomes were therefore analyzed as complementary measures of participant-experienced neuropathy symptom burden.

### Chart Abstraction for Baseline Lymphoma Stage and Glycemic Status

Baseline lymphoma stage and glycemic status at the time of diagnosis or treatment initiation were abstracted from the electronic medical record. HbA1c was recorded using the value closest to lymphoma diagnosis or treatment baseline when available. When HbA1c was unavailable, the glucose value closest to the diagnosis was recorded and used to calculate a glucose-derived estimate of HbA1c [36]. While measured HbA1c reflects average glycemia over the preceding 2-3 months, HbA1c values estimated from contemporaneous glucose measurements may be disproportionately influenced by short-term hyperglycemia, including glucocorticoid-associated hyperglycemia around chemotherapy initiation [37, 38].

## Analysis

### Sample Size Determination

Enrollment of 25 eligible participants was intended to assess feasibility of clinically aligned serial NfL and patient-reported outcome collection and to generate preliminary estimates for future powered studies. Exploratory biomarker-outcome analyses were interpreted descriptively.

### Feasibility assessment

Feasibility was assessed using prespecified criteria intended to inform the design of a larger longitudinal study. First, collection of an early-treatment assessment was considered feasible if blood and clinical outcome data were obtained from more than 75% of enrolled participants either at T1/Pre-C1 or, when pretreatment collection was not possible because of clinical workflow or later recruitment, at T2/Pre-C2. This criterion was selected to reflect the practical challenges of embedding biospecimen and patient-reported outcome collection into routine NHL care. Second, the rate of retention of participants who remained eligible for study inclusion at T4/Pre-C6 was calculated, and prospective assessment of this final on-treatment timepoint was considered feasible if data collection was achieved among at least 50% of eligible enrollees [39]. This criterion was designed to (a) inform estimates of enrollee attrition due to NHL disease progression between T1 and T4, meaning disease worsening that caused disenrollment from the study due to life-saving updates to the treatment plan, and (b) assess attrition rate among enrollees who remained eligible for study participation. Third, completion of longitudinal assessments was considered feasible if planned treatment-period assessments were collected within the anticipated chemotherapy observation window, allowing for clinically necessary rescheduling of visits due to illness, treatment delays, hospitalization, or other factors. This criterion was designed to evaluate whether clinically aligned longitudinal sampling could be implemented within routine NHL care and to inform allowable timepoint collection windows for future larger studies. We assessed feasibility across these three criteria, requiring that all criteria be deemed feasible to declare the study feasible.

### Statistical Analysis

Demographic and clinical characteristics were summarized using descriptive statistics. Continuous variables were summarized using means, medians, standard deviations, minima, and maxima; categorical variables were summarized using frequencies and percentages. Participant codes shown in figures and supplementary tables are randomized public analysis codes generated for this report and do not correspond to study IDs or clinical identifiers. Because plasma NfL concentrations were right-skewed, NfL was analyzed on the log10 scale. The primary longitudinal analysis used a linear mixed-effects model with log10-transformed NfL as the outcome, timepoint as a fixed effect, and participant-level random intercepts to account for repeated measures. Analytical sample sizes differed by observed data availability, and no imputation was performed for any analysis. Model estimates were back-transformed and presented as model-estimated geometric means (pg/mL) with 95% confidence intervals. Relative changes from T1/Pre-C1 were calculated from the model-estimated geometric means. An exploratory regimen model included chemotherapy regimen and timepoint-by-regimen interaction to compare CHOP and EPOCH treatment groups.

CIPN20 was analyzed using a linear mixed-effects model with log-10 transformed raw CIPN20 total score as the outcome, timepoint as a fixed effect, and participant-level random intercepts. Estimates were back-transformed for display, and model-based estimated marginal mean contrasts compared each later timepoint with T1/Pre-C1. BPI-SF severity scores and CTCAE neuropathy grades were summarized descriptively and evaluated using fixed effects for timepoint; bootstrap confidence intervals and p-values were used because model residuals did not meet the assumptions of normality. T5/Post patient-reported outcome data were analyzed as exploratory long-term follow-up data and displayed separately from the treatment-period visit sequence. T5/Post CTCAE data were descriptive only because only one CTCAE observation was available.

Exploratory associations between plasma NfL and CIPN20 scores were evaluated using two-sided Spearman rank correlations because of the small sample sizes and the descriptive, nonparametric nature of these analyses. For the pooled visit-level analysis, all paired treatment-period observations with both plasma NfL and CIPN20 data were included and analyzed descriptively; repeated observations from the same participant were not treated as independent evidence for prediction. Exploratory lagged analyses evaluated whether early within-person change in log10 plasma NfL from the first available early-treatment sample to T3/Pre-C4 was associated with subsequent change in CIPN20 total score from T3/Pre-C4 to T4/Pre-C6. Post hoc sensitivity analyses evaluated early T2/Pre-C2 log10 plasma NfL in relation to later CIPN20 total score at T3/Pre-C4 and T4/Pre-C6. Squared Spearman rank correlations (ρ²) were displayed descriptively in selected panels to summarize rank-based monotonic association. Least-squares regression lines were overlaid as visual reference.

Exploratory vincristine dose-modification analyses compared participants with no vincristine dose modification versus any vincristine dose modification by T4/Pre-C6. Changes in log10 plasma NfL and CIPN20 total score from baseline to T4/Pre-C6 were compared between groups using Mann-Whitney U tests. All analyses were interpreted in the context of the study’s feasibility-focused design and limited sample size.

## Results

### Cohort and baseline characteristics

Of the 28 individuals who underwent screening, 25 consented and enrolled (Fig. 2A). The baseline descriptive cohort included 25 participants, of whom 16 were treated with CHOP and 9 with EPOCH (Table 1). Baseline demographic and clinical characteristics, including age, sex, race/ethnicity, ECOG performance, lymphoma stage, and glycemic status, are summarized in Table 1. Sixteen were treated with CHOP, which was administered in the outpatient setting, typically during an approximately 1.5-hour infusion visit. Nine were treated with EPOCH, which typically required inpatient admission for approximately 5 days and included continuous infusion over 96 hours. Participants receiving CHOP were, on average, older than those receiving EPOCH (mean 70.1 vs 53.0 years), more likely to be male (n = 9, 56.3%), and more likely to be White (n = 14, 87.5%). Stage was documented for 18 of 25 participants; among staged participants, 16 had stage III/IV or III-IV disease and 2 had stage II/IIA disease, while 7 were unstaged or not documented (ND) in the extracted record. Measured HbA1c was available for 19 participants, and glucose-derived estimated HbA1c was available for 6 additional participants. The combined measured/estimated HbA1c median was 5.5% (range, 4.2-8.5); 6 participants had values in the diabetes range (≥6.5%, all from CHOP). No participants had concurrent autoimmune disease or were taking a CYP3A4 inhibitor, a potential interactor with vincristine [40, 41], at baseline. One participant with human immunodeficiency virus (HIV) was included in the primary analysis because our previous retrospective study showed no association between HIV and CIPN [42]. Separately, one participant was identified as a high symptom-severity outlier (i.e., ≥2 standard deviations above the mean), and we therefore also report a post hoc sensitivity analysis in which this outlier was removed.

**Figure 2.**
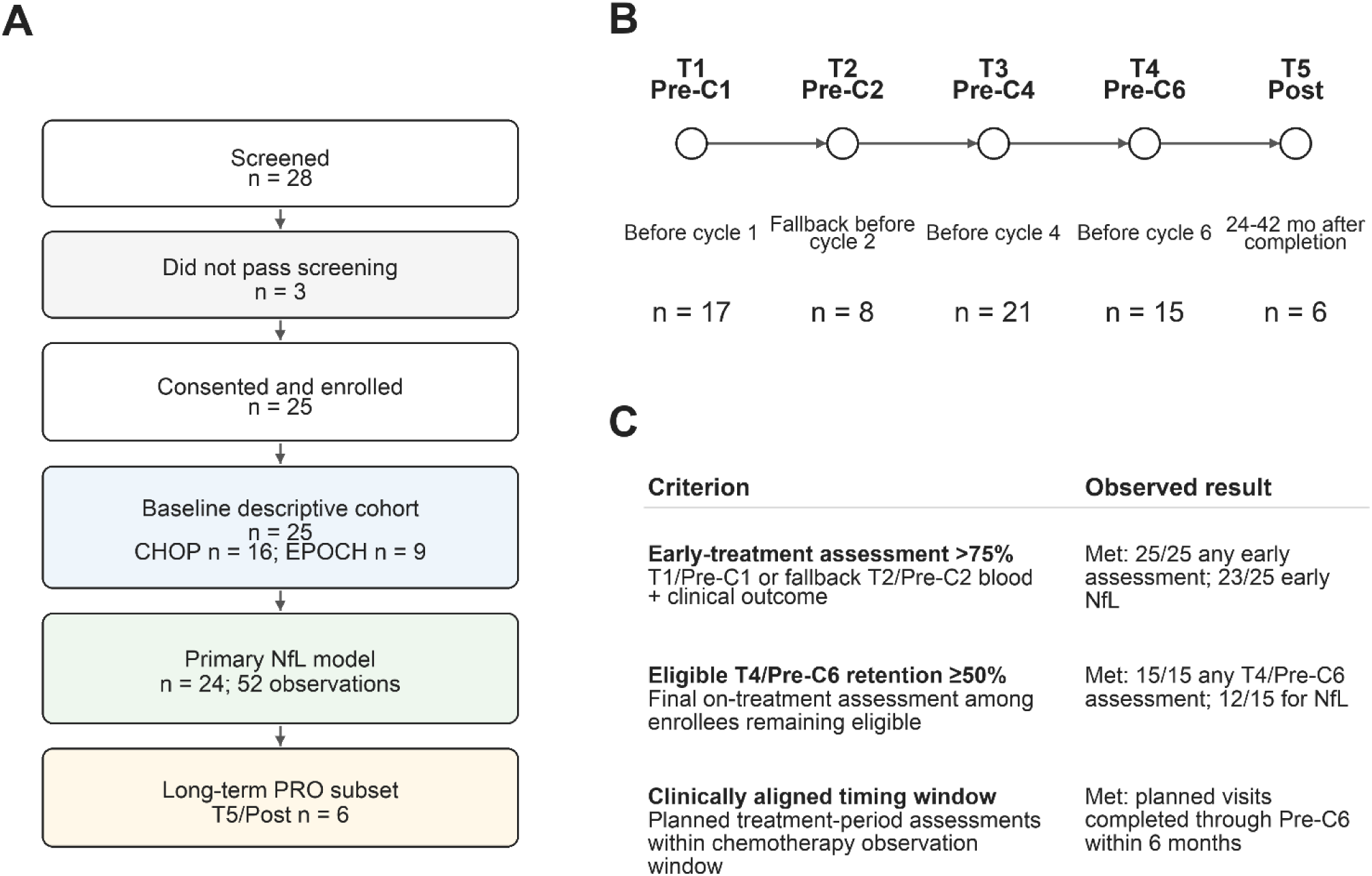
Overview of study execution and feasibility. (A) STROBE (Strengthening the Reporting of Observational Studies in Epidemiology) cohort flow from screening through analytic subsets. Twenty-eight patients were screened; 25 eligible participants contributed to the baseline descriptive cohort. The primary treatment-period plasma NfL model included 24 participants with at least one valid treatment-period plasma NfL measurement, contributing to 52 NfL observations, and the long-term T5/Post PRO subset included 6 participants. (B) Completed assessments by timepoint. Numbers indicate available assessments at each visit: T1/Pre-C1 n = 17, T2/Pre-C2 n = 8, T3/Pre-C4 n = 21, T4/Pre-C6 n = 15, and T5/Post n = 6. (C) Feasibility criteria and observed results. The early assessment criterion was met (25/25 participants had any assessment; 23/25 had early plasma NfL). Retention through T4/Pre-C6 was met (15/15 had any assessment; 14/15 had PRO data; 12/15 had NfL data). The clinically aligned treatment-window criterion was met, with planned visits completed through T4/Pre-C6 within 6 months.

**Table 1.** Baseline demographic and clinical characteristics by treatment regimen. Baseline demographic and clinical characteristics of the eligible descriptive cohort, stratified by chemotherapy regimen. Continuous variables are summarized as mean (SD) and range or median (range), as indicated. Categorical variables are summarized as n (%). Cancer stage reflects chart-documented stage; unstaged/not documented (ND) indicates no stage information in the extracted record. HbA1c/eA1c combines measured HbA1c with glucose-derived estimated HbA1c when HbA1c was unavailable; glucose-derived eA1c values were used descriptively only. CHOP, cyclophosphamide, doxorubicin, vincristine, and prednisone; EPOCH, etoposide, prednisone, vincristine, cyclophosphamide, and doxorubicin; ECOG, Eastern Cooperative Oncology Group; BMI, body mass index. HbA1c, hemoglobin A1c; eA1c, estimated HbA1c.

| Characteristic | CHOP (n = 16) | EPOCH (n = 9) | Total sample (n = 25) |
| --- | --- | --- | --- |
| Age, mean (SD); range | 70.1 (13.4); 31-87 | 53.0 (21.2); 19-74 | 63.4 (18.4); 19-87 |
| Sex, n (%) |  |  |  |
| *Male* | 9 (56.3) | 3 (33.3) | 12 (48.0) |
| *Female* | 7 (43.8) | 6 (66.7) | 13 (52.0) |
| Race/ethnicity, n (%) |  |  |  |
| *White* | 14 (87.5) | 6 (66.7) | 20 (80.0) |
| *Non-White* | 2 (12.5) | 3 (33.3) | 5 (20.0) |
| *Black* | 2 (12.5) | 1 (11.1) | 3 (12.0) |
| *Asian* | 0 (0.0) | 0 (0.0) | 0 (0.0) |
| *Hispanic* | 0 (0.0) | 1 (11.1) | 1 (4.0) |
| *More than 1 race/ethnicity* | 0 (0.0) | 1 (11.1) | 1 (4.0) |
| ECOG performance status, n (%) |  |  |  |
| *0* | 9 (56.3) | 3 (33.3) | 12 (48.0) |
| *1* | 4 (25.0) | 2 (22.2) | 6 (24.0) |
| *2* | 3 (18.8) | 1 (11.1) | 4 (16.0) |
| *Missing* | 0 (0.0) | 3 (33.3) | 3 (12.0) |
| BMI, median (range) | 29.0 (19.8-45.6) | 25.9 (20.0-38.5) | 27.5 (19.8-45.6) |
| Diabetes, yes, n (%) | 5 (31.3) | 1 (11.1) | 6 (24.0) |
| Depression medication, yes, n (%) | 5 (31.3) | 0 (0.0) | 5 (20.0) |
| Cancer stage at baseline, n (%) |  |  |  |
| *Stage II/IIA* | 0 (0.0) | 2 (22.2) | 2 (8.0) |
| *Stage III/IV* | 13 (81.3) | 3 (33.3) | 16 (64.0) |
| *Unstaged/not documented* | 3 (18.8) | 4 (44.4) | 7 (28.0) |
| HbA1c/eA1c, median (range), % | 6.1 (4.7-8.5) | 5.4 (4.2-6.0) | 5.5 (4.2-8.5) |
| HbA1c/eA1c source, n (%) |  |  |  |
| *Measured HbA1c* | 12 (75.0) | 7 (77.8) | 19 (76.0) |
| *Glucose-derived eA1c* | 4 (25.0) | 2 (22.2) | 6 (24.0) |
| HbA1c/eA1c category, n (%) |  |  |  |
| *Normal (<5.7%)* | 6 (37.5) | 8 (88.9) | 14 (56.0) |
| *Prediabetes range (5.7-6.4%)* | 4 (25.0) | 1 (11.1) | 5 (20.0) |
| *Diabetes range (≥6.5%)* | 6 (37.5) | 0 (0.0) | 6 (24.0) |

### Feasibility criteria were met

All prespecified feasibility criteria were met (**Fig. 2B, C**). Early-treatment data collection exceeded the prespecified threshold, with 25 of 25 participants (100%) contributing patient and clinician assessment results at either T1/Pre-C1 or T2/Pre-C2 and 23 of 25 (92%) contributing early NfL data. Retention through the final planned on-treatment assessment, T4/Pre-C6, was achieved for 15 of 15 (100%) for any assessments, while T4/Pre-C6 NfL collection was achieved for 12 of 15 participants (80%). Planned on-treatment assessments were collected at clinically aligned chemotherapy visits through T4/Pre-C6, corresponding to an observation window of up to 6 months. These findings support the operational feasibility of clinically aligned longitudinal sampling in routine lymphoma care, while also identifying NfL-specific retention at later treatment timepoints as an important consideration for future studies.

### Plasma NfL increased early and remained elevated during treatment

Raw NfL distributions were skewed, supporting log10-transformed modeling. In the primary longitudinal mixed-effects model accounting for repeated measures within participants, the model-estimated plasma NfL increased from 26.3 pg/mL at T1/Pre-C1 (95% CI, 18.9-36.8) to 91.6 pg/mL at T2/Pre-C2 (95% CI, 59.5-141.2), 107.2 pg/mL at T3/Pre-C4 (95% CI, 77.3-148.7), and 94.3 pg/mL at T4/Pre-C6 (95% CI, 66.1-134.4). Relative to T1/Pre-C1, this corresponded to increases of 248.0% at Pre C2, 307.4% at T3/Pre-C4, and 258.2% at T4/Pre-C6 (all p < 0.001) (**Fig. 3A, B; Fig. S1; Table 2**). This pattern is consistent with an early treatment-associated increase in plasma NfL followed by sustained elevation through later on-treatment timepoints, with the highest estimated geometric mean observed at T3/Pre-C4 and persistent elevation at T4/Pre-C6. Because some participants first entered the study at T2/Pre-C2, and T1/Pre-C1 and T2/Pre-C2 values were not available from the same complete set of participants, this contrast should be interpreted as an early collection-timing comparison in the context of heterogeneous baseline timing, rather than uniform within-person pretreatment-to-postexposure change.

**Figure 3.**
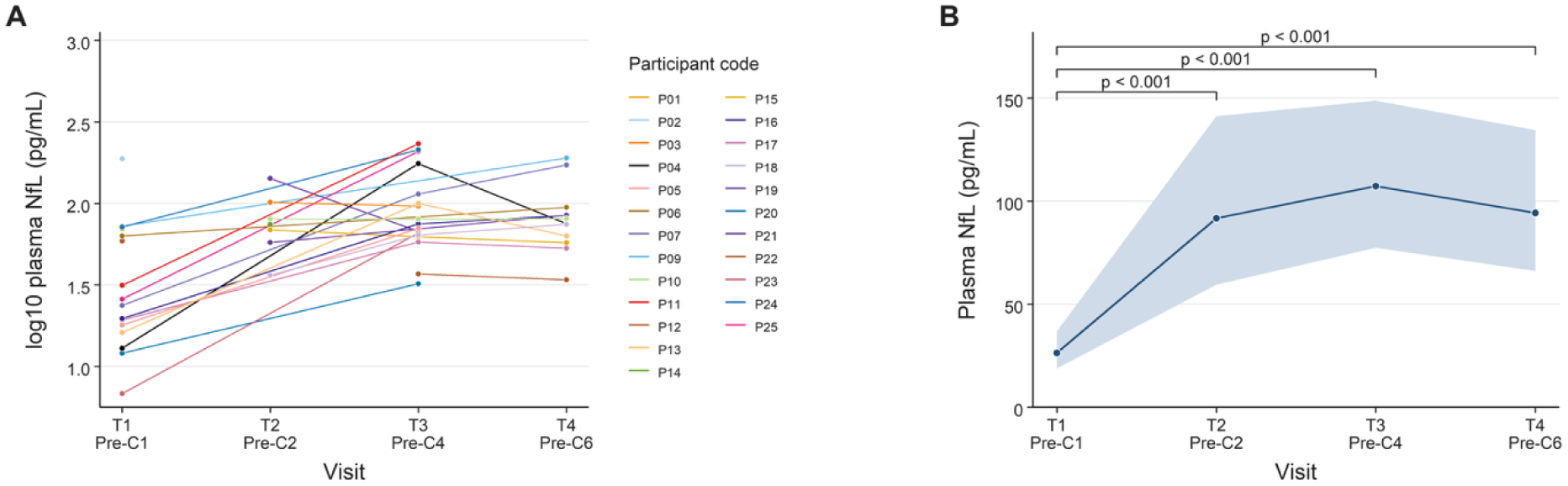
Longitudinal plasma NfL during vincristine-containing chemotherapy. (A) Individual participant plasma NfL trajectories shown on the log10 scale across treatment-period visits. Lines connect repeated observations within participants. (B) Model-estimated plasma NfL geometric means and 95% confidence intervals, back-transformed to pg/mL from the primary mixed-effects model fit on log10-transformed NfL. The model included timepoint as a fixed effect and a participant-level random intercept. Points show back-transformed model estimates and the shaded/error interval shows the 95% confidence interval. P values shown in the panel are model-based estimated marginal mean contrast p values comparing each follow-up timepoint with T1/Pre-C1. Compared with T1/Pre-C1, plasma NfL was higher at T2/Pre-C2 (p = 4.16e-06) T3/Pre-C4 (p = 6.41e-10) and T4/Pre-C6 (p = 3.39e-08). NfL, neurofilament light chain.

**Table 2.**
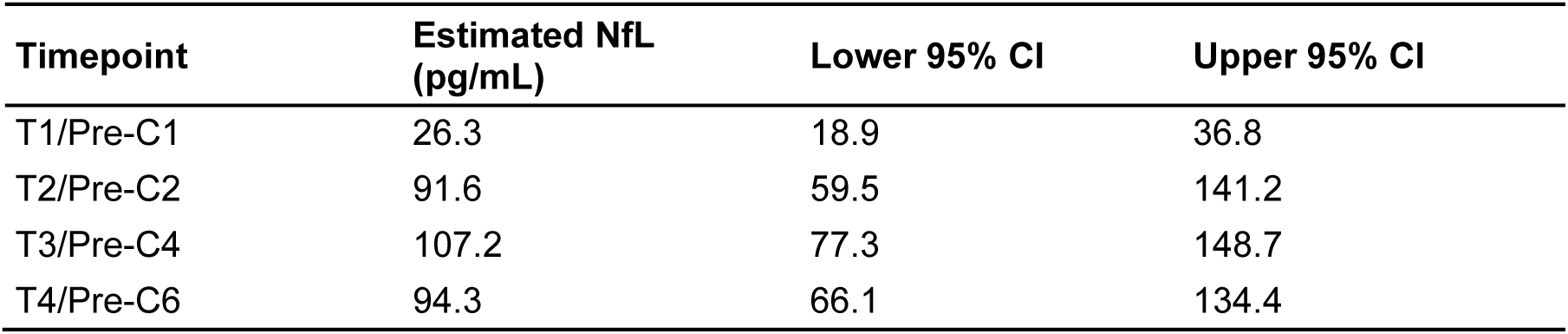
Model-estimated plasma NfL concentrations from the primary mixed-effects model. Plasma NfL was analyzed on the log10 scale using a mixed-effects model with timepoint as a fixed effect and participant-level random intercept. Estimates and 95% confidence intervals were back-transformed and are presented as geometric mean plasma NfL concentrations in pg/mL. NfL, neurofilament light chain; CI, confidence interval.

A secondary model including the chemotherapy regimen and a timepoint-by-regimen interaction did not identify significant differences between CHOP and EPOCH in this pilot study. The main effect for EPOCH was not significant compared to CHOP (p = 0.779), and timepoint-by-regimen interaction terms were not significant at T2/Pre-C2 (p = 0.132), T3/Pre-C4 (p = 0.929), or T4/Pre-C6 (p = 0.356) compared to T1/Pre-C1 (**Fig. S2**).

### Patient-reported and clinical neuropathy measures increased during chemotherapy

CIPN20 scores increased with chemotherapy exposure. In the mixed-effects model, model-estimated geometric mean CIPN20 was 20.9 at T1/Pre-C1 (95% CI, 19.1-22.8), 20.8 at T2/Pre-C2 (95% CI, 18.3-23.6), 23.2 at T3/Pre-C4 (95% CI, 21.4-25.1), 25.8 at T4/Pre-C6 (95% CI, 23.4-28.4), and 24.5 at >2 years post chemotherapy completion (Post; 95% CI, 21.2-28.4). Relative to T1/Pre-C1, this corresponded to a 23.4% increase at T4/Pre-C6 (95% CI, 9.1-39.7; p = 0.001). Changes were smaller at T3/Pre-C4 (10.9% increase; 95% CI, -0.7 to 23.8; p = 0.070) and Post (17.5% increase; 95% CI, -0.2 to 38.4; p = 0.053), while T2/Pre-C2 was essentially unchanged from T1/Pre-C1 (-0.6%; p = 0.900) (**Fig. 4A**; **Table 3**).

**Figure 4.**
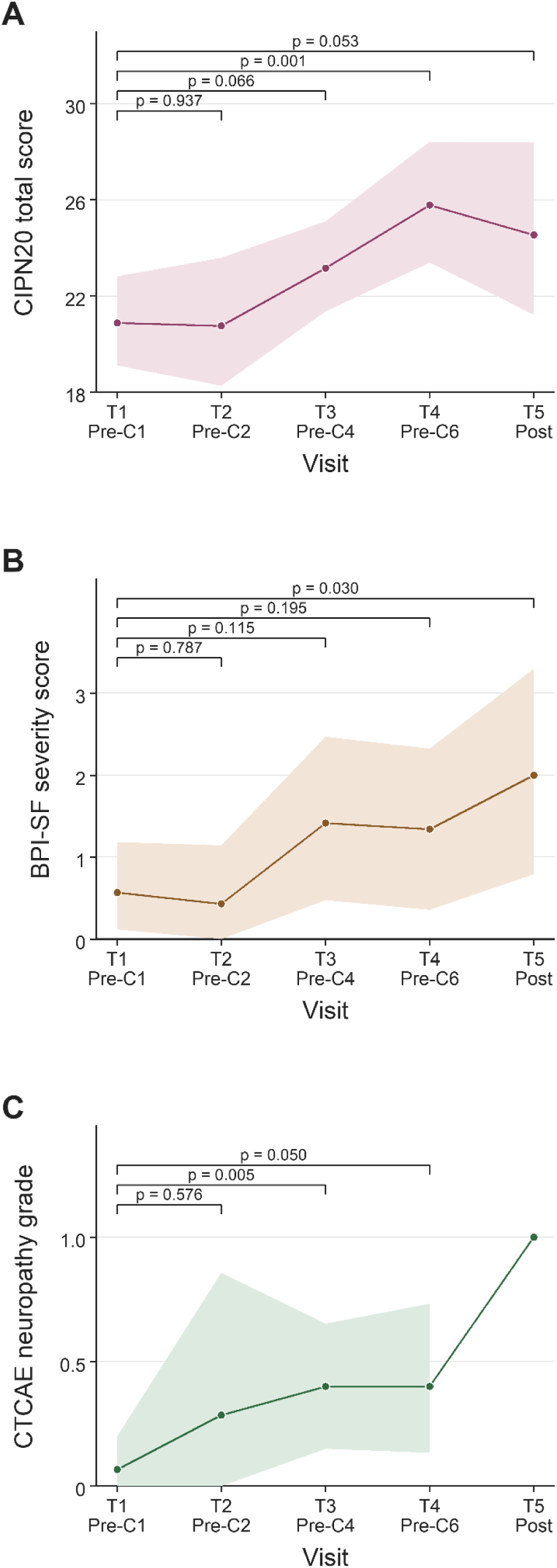
Patient-reported and clinician-graded outcomes over time. (A) Model-estimated EORTC QLQ-CIPN20 total score (retained raw calculated total) from the marginal timepoint model output. (B) Model-estimated BPI-SF severity score from the marginal timepoint model output. (C) Model-estimated CTCAE neuropathy grade from the marginal timepoint model output. The confidence limits below 0 were truncated at 0 for display. (A)-(C) shaded bands show treatment-period estimated marginal means and 95% confidence intervals from T1/Pre-C1 through exploratory T5/Post estimated means and 95% confidence intervals where estimable. Model-based estimated marginal mean contrasts comparing each later timepoint with T1/Pre-C1, shown as p values. T5/Post CTCAE was descriptive only because only one observation was available. CIPN20, Chemotherapy-Induced Peripheral Neuropathy 20-item questionnaire; BPI-SF, Brief Pain Inventory-Short Form; CTCAE, Common Terminology Criteria for Adverse Events.

**Table 3.** Model-estimated patient-reported and clinician-graded outcomes by timepoint. CIPN20 values represent model-estimated back-transformed raw CIPN20 total scores. CTCAE neuropathy grade and BPI-SF severity scores are presented as descriptive model-estimated means with 95% confidence intervals. The T5/Post CTCAE value reflects one available observation and was not used for inferential analysis. CIPN20, Chemotherapy-Induced Peripheral Neuropathy questionnaire; CTCAE, Common Terminology Criteria for Adverse Events; BPI-SF, Brief Pain Inventory-Short Form; CI, confidence interval.

| Timepoint | CIPN20 geometric mean (95% CI) | CTCAE mean (95% CI) | BPI-SF pain severity mean (95% CI) |
| --- | --- | --- | --- |
| T1/Pre-C1 | 20.9 (19.1-22.8) | 0.1 (0.0-0.2) | 0.6 (0.1-1.2) |
| T2/Pre-C2 | 20.8 (18.3-23.6) | 0.3 (0.0-0.9) | 0.4 (0.0-1.1) |
| T3/Pre-C4 | 23.2 (21.4-25.1) | 0.4 (0.1-0.7) | 1.4 (0.5-2.5) |
| T4/Pre-C6 | 25.8 (23.4-28.4) | 0.4 (0.1-0.7) | 1.3 (0.4-2.3) |
| T5/Post | 24.5 (21.2-28.4) | 1.0 (n = 1; no CI) | 2.0 (0.8-3.3) |

BPI severity scores and clinician-recorded CTCAE toxicity assessment provided additional clinical context and were interpreted descriptively given sparse counts, routine-care ascertainment, and limited sample size. BPI severity scores were generally low during treatment, with mean scores ranging from 0.43 to 1.42 across pre-cycle treatment visits and 2.0 among participants with T5/Post follow-up data (**Fig. 4B**; **Table 3**).

CTCAE-defined grade ≥1 neuropathy was present in 5 of 15 participants (33.3%) at T4/Pre-C6, including grade 1 neuropathy in three participants and grade 2 neuropathy in two participants. Raw mean CTCAE grade was 0.07 at T1/Pre-C1 (n = 15), 0.29 at T2/Pre-C2 (n = 7), 0.40 at T3/Pre-C4 (n = 20), 0.40 at T4/Pre-C6 (n = 15), and 1.0 at T5/Post (n = 1). In the exploratory CTCAE timepoint model, estimated mean CTCAE grade was 0.1 at T1/Pre-C1 (95% CI, 0.0-0.2), 0.3 at T2/Pre-C2 (95% CI, 0.0-0.9), 0.4 at T3/Pre-C4 (95% CI, 0.1-0.7), and 0.4 at T4/Pre-C6 (95% CI, 0.1-0.7). Absolute increases from T1/Pre-C1 were 0.2 at T2/Pre-C2 (95% CI, -0.1 to 0.8; p = 0.600), 0.3 at T3/Pre-C4 (95% CI, 0.1-0.6; p = 0.005), and 0.3 at T4/Pre-C6 (95% CI, 0.0-0.7; p = 0.050) (**Fig. 4C**; **Table 3**).

### Exploratory NfL-CIPN20 analyses suggested group-level association but limited individual-level concordance

Because both plasma NfL and neuropathy outcomes increased over treatment, we next explored whether higher NfL was associated with greater patient-reported symptom burden at the visit level or whether earlier NfL change predicted subsequent CIPN20 worsening. In exploratory pooled visit-level analyses combining all paired NfL and CIPN20 observations across treatment timepoints, higher log10 NfL was modestly associated with higher CIPN20 scores (n = 51; Spearman ρ = 0.393, p = 0.004). Timepoint-specific relationships were inconsistent and limited by small sample sizes (**Fig. 5A; Table S1**), and exploratory lagged within-person analyses did not show that early NfL change predicted subsequent worsening of CIPN20. Early change in log10 NfL from the first available early-treatment sample to T3/Pre-C4 was not associated with later change in CIPN20 from T3/Pre-C4 to T4/Pre-C6 (n = 10; Spearman ρ = 0.04, p = 0.91) (**Fig. 5B**).

**Figure 5.**
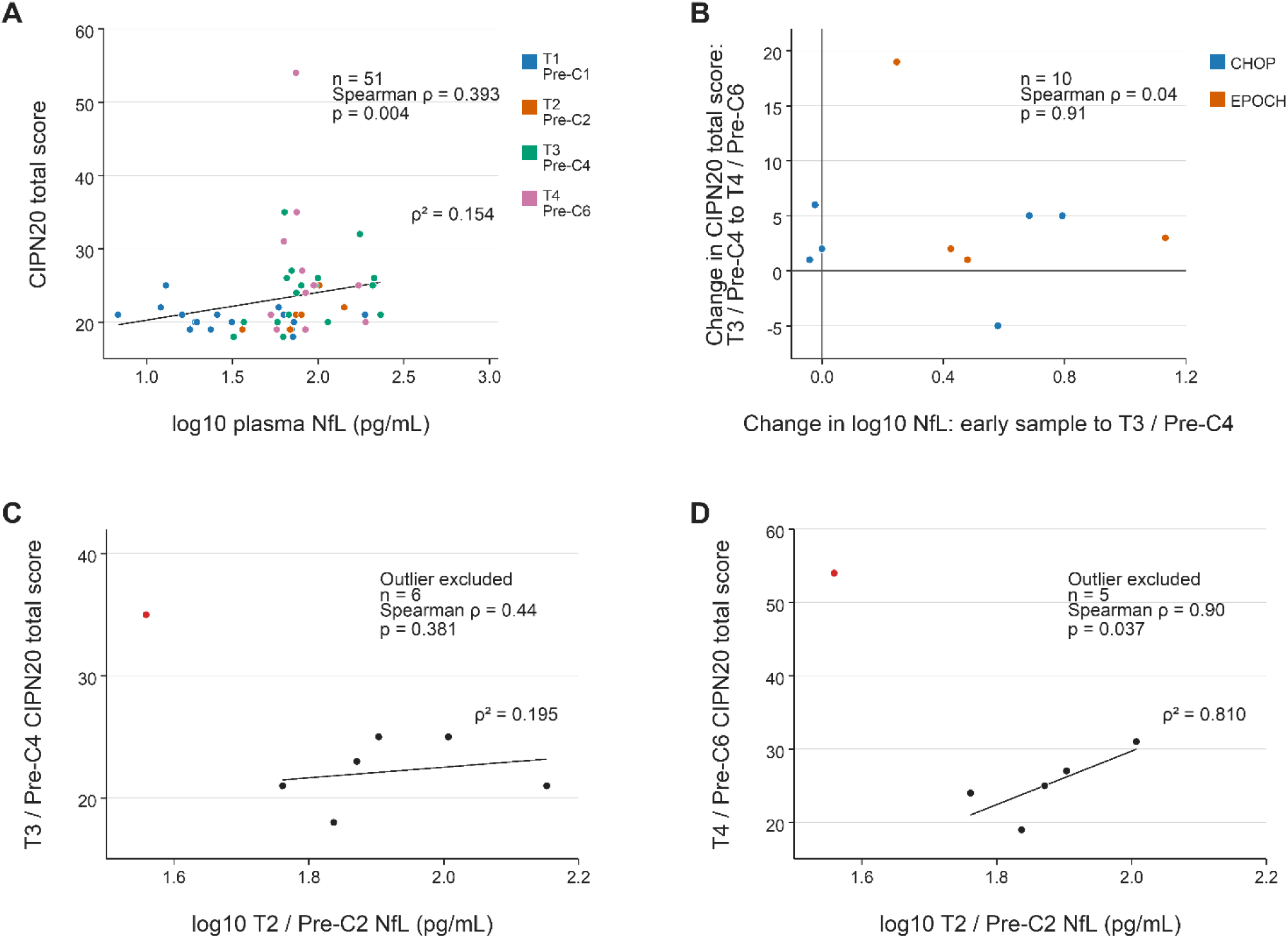
Exploratory relationships between plasma NfL and patient-reported neuropathy, including sensitivity analyses. (A) Pooled visit-level association between log10 plasma NfL and CIPN20 total score across treatment-period visits with paired measurements. Points are colored by visit. Association was tested using a two-sided Spearman rank correlation: ρ = 0.393, p = 0.004, n = 51. The displayed ρ² value represents the squared Spearman correlation. (B) Exploratory lagged within-person analysis comparing change in log10 plasma NfL from the first available early-treatment sample to T3/Pre-C4 with subsequent change in CIPN20 total score from T3/Pre-C4 to T4/Pre-C6. Points are colored by chemotherapy regimen; zero reference lines indicate no change. Association was tested using a two-sided Spearman rank correlation: ρ = 0.04, p = 0.91, n = 10. (C, D) Post hoc sensitivity analyses examining early T2/Pre-C2 log10 plasma NfL in relation to later CIPN20 total score at (C) T3/Pre-C4 and (D) T4/Pre-C6. The high symptom-severity outlier (≥2 standard deviations above the mean) is highlighted in red but not labeled. Black lines show least-squares linear fits after excluding the highlighted outlier and are included only as visual references. In-panel statistics show two-sided Spearman rank correlations after excluding the highlighted outlier: (C) ρ = 0.441, p = 0.381, n = 6; (D) ρ = 0.900, p = 0.037, n = 5. Displayed ρ² values represent squared Spearman correlations. With the highlighted outlier included, the corresponding Spearman correlations were (C) ρ = -0.109, p = 0.816, n = 7, and (D) ρ = 0.086, p = 0.872, n = 6. These post hoc analyses are exploratory and underpowered and should be interpreted as evidence of individual-level heterogeneity rather than validation of a predictive NfL-CIPN20 relationship. NfL, neurofilament light chain; CIPN20, Chemotherapy-Induced Peripheral Neuropathy questionnaire; CHOP, cyclophosphamide, doxorubicin, vincristine, and prednisone; EPOCH, etoposide, prednisone, vincristine, cyclophosphamide, and doxorubicin.

A post hoc sensitivity analysis further examined the relationship between early NfL and later CIPN20. We excluded one data point determined to be an outlier, because of a symptom severity score more than 2 standard deviations outside the mean and a biologically justified reason for exclusion (i.e., additional disease burden associated with immunodeficiency), the association between T2/Pre-C2 NfL and T4/Pre-C6 CIPN20 became positive by rank correlation, but the analysis remained based on very small numbers and should be interpreted only as hypothesis-generating. This sensitivity analysis supports the presence of individual-level heterogeneity and reinforces that plasma NfL and CIPN20 should not be treated as interchangeable measures in this pilot dataset (Fig. 5C, 5D).

### Dose modifications occurred for a quarter of participants

Vincristine dose modifications – including reductions, holds, and discontinuations – occurred for 6 out of 25 (24%) of the cohort with reasons that included neuropathy, ileus, constipation, and multifactorial clinical judgment (**Table S2**). No effect of dose modification was detected (**Fig. S3A, S3B**). As depicted in Fig. S3A, however, dose-modified participants had a numerically larger median increase in log10 NfL from baseline than participants without dose modification by T4/Pre-C6, (median Δlog10 NfL, 0.632 vs 0.244; approximately 4.3-fold vs 1.75-fold increase; Mann-Whitney p = 0.194). In contrast, median CIPN20 change from baseline to T4/Pre-C6 was lower in the dose-modified group than in the no-dose-modification group (median ΔCIPN20, 0 vs 6; Mann-Whitney p = 0.138). These exploratory comparisons did not meet conventional statistical significance and were limited by small sample size, heterogeneous timing, and heterogeneous reasons for dose modification. However, they suggest that the delivery of vincristine and dose modification may alter the observed relationship between NfL kinetics and patient-reported symptom trajectories.

### Exploratory long-term patient-reported outcomes suggested CIPN persistence

Long-term patient-reported outcome data were available for six survivors 24-42 months after completion of vincristine-based chemotherapy. At T5/Post, raw CIPN20 total scores ranged from 20 to 36, and BPI-SF worst-pain item ranged from 0 to 4.25. The two participants with the highest T5/Post CIPN20 scores also had the highest T5/Post BPI severity scores as well as vincristine dose modification during treatment. It is important to note, however, dose modifications cannot be assumed to have occurred because of neuropathy worsening. Indeed, dose modifications did not lead universally to high T5/Post symptom burden (**Table S3**).

Complete treatment-period NfL values from the first available early-treatment sample through T3/Pre-C4 and T4/Pre-C6 were available for four participants. Given the small number of participants with complete NfL trajectories and long-term PRO data, these long-term data are presented descriptively to show that patient-reported neuropathy symptoms can persist years after therapy and that real-world dose modification is an important interpretive variable when evaluating NfL and symptom trajectories. Further studies are required to test the hypothesis that a specific NfL trajectory shape, dose reduction, and functional outcome measures can serve as a multidomain risk model for persistent neuropathy.

## Discussion

Feasibility criteria were met, and our hypothesis that plasma NfL would increase with vincristine exposure was upheld. We were able to collect serial plasma NfL within the course of routine care, administer neuropathy (CIPN20) and pain (BPI) patient-reported symptom assessments at the point of care or via emailed surveys, and abstracted clinician-recorded CTCAE neuropathy grades, vincristine dose-modification information, and relevant baseline information from the medical chart review. The resulting cohort data demonstrate that plasma NfL increased after initial vincristine exposure and remained elevated across later chemotherapy infusion cycles. Exploratory analyses of the relationship between NfL and CIPN revealed that the measures increased together when timepoints were pooled, however, at individual timepoints, NfL and CIPN20 outcomes showed no correlation within this small dataset. The post-hoc sensitivity analysis suggested that early NfL changes may predict later CIPN20 changes. These findings support further study of plasma NfL as a potential biomarker of CIPN and related symptom burden resulting from vincristine-containing lymphoma chemotherapy.

Mechanistically, our interpretation that plasma NfL concentrations are sensitive to vincristine exposure is plausible. A preclinical rodent study indicates increased NfL following vincristine treatment, further supporting the biological plausibility of a vincristine-associated NfL response consistent with vincristine-associated distal axonal injury [22]. Similarly, our results showed an overall increase in NfL, yet this increase did not show a simple monotonic cumulative-dose pattern typically reported for other chemotherapeutic agents such as platinum and paclitaxel [15, 17–21, 23]. Our data are in line with a long-term follow-up study of vincristine-treated participants with hematological cancer, as well as with early-onset vincristine-induced peripheral neuropathy in B-cell lymphoma, which showed no association with the total vincristine dose or the number of treatment cycles [3, 4]. Together, these findings support the notion of a potentially distinct biology underlying vincristine-induced NfL release and neurodegeneration that contribute to CIPN.

Emerging mechanistic data suggest that NfL release may also intersect with injury-associated immune responses. A recent study reported that NfL released after neuronal damage can activate myeloid cells and promote neuroinflammatory responses in a model of amyotrophic lateral sclerosis [43]. This may be relevant to vincristine-associated neurotoxicity because vincristine is increasingly understood to involve immune-mediated processes in preclinical studies, including neuroimmune signaling, astrocyte activation, macrophage/myeloid responses, cytokine release, and pain sensitization [44–49]. Although our study was not designed to test whether NfL contributes functionally to neuroinflammation in CIPN, these observations motivate investigating plasma NfL in the context of a broader injury and immune response rather than as a same-day symptom-severity readout.

Immune modulation of vincristine-induced neurotoxic injury may help explain why NfL and CIPN20 were related at the pooled-visit level but showed limited concordance at the individual level. Patient-perceived symptoms may emerge before, during, or after detectable NfL release, reflecting sensory nerve dysfunction, altered excitability, immune activation, pain sensitization, and functional adaptation. Conversely, circulating NfL may remain elevated after the peak of acute symptom worsening if neuroaxonal injury, clearance kinetics, or neuroimmune responses persist. The post hoc sensitivity analysis, further supports the possibility of individual-level heterogeneity. However, this analysis was underpowered and hypothesis-generating; it should not be interpreted as evidence of a predictive NfL-CIPN20 relationship. Rather, it reinforces the more conservative conclusion that NfL and CIPN20 are complementary but non-interchangeable measures.

Clinician-recorded CTCAE neuropathy and patient-reported CIPN20 should not be expected to align perfectly because routine-care CTCAE grading reflects clinician-documented toxicity during clinical encounters [35], whereas CIPN20 captures symptom burden through a structured patient-reported instrument [30, 31]. In this cohort, CIPN20 showed a clearer longitudinal increase in patient-reported symptoms than clinician-graded CTCAE neuropathy scores, consistent with prior studies indicating that clinician- and patient-reported CIPN measures are related but not interchangeable [50–53]. Similar temporal divergence between clinician-assessed neuropathy and patient-reported burden has also been reported in the NOVIT study of hematologic patients receiving vincristine-, bortezomib-, or lenalidomide-containing therapy [54]. Consistent with this broader literature, Andersen et al. (2024) emphasized that NfL reflects neuroaxonal injury and should be interpreted alongside clinical and patient-reported outcomes rather than as a direct readout of symptom severity [15]. Together, these observations support multi-modal assessment and reinforce that NfL, clinician grading, and PROs provide complementary rather than redundant information.

The BPI severity provided additional clinical context but should not be interpreted as a CIPN-specific endpoint, as it captures overall pain intensity from cancer, treatment, comorbidities, musculoskeletal conditions, or other causes. In this cohort, BPI severity scores were generally low during treatment, whereas CIPN20 scores increased. This pattern is consistent with the idea that patient-reported neuropathy burden may include numbness, tingling, sensory change, motor symptoms, and functional impact that are not fully captured by a general pain severity measure.

Several limitations should be emphasized. First, this was a small, single-center pilot study with substantial missingness for PROs, clinician-recorded CTCAE toxicity grading, BPI severity score, dose-modification variables, and complete longitudinal NfL trajectories. Second, early sampling was heterogeneous because clinically aligned collection in routine lymphoma care did not always permit T1/Pre-C1 sampling before first vincristine exposure; therefore, T2/Pre-C2 served as a protocol-defined fallback early-treatment window. This limits interpretation of early timepoint contrasts and reinforces the need for standardized pretreatment sampling in future studies. Third, NfL is not specific to CIPN and can be influenced by age, renal function, body size/distribution volume, neurological comorbidity, and assay factors [13–15, 24–27]. These factors may contribute to between-participant variability, particularly in a small cohort, and support interpreting NfL longitudinally rather than as a cross-sectional diagnostic threshold. Fourth, clinician assessment was limited to standard-of-care CTCAE toxicity documentation. The study did not administer CTCAE questions directly, and toxicity grading in routine practice is subject to provider-level variability, particularly for symptoms such as paresthesia that rely on clinical judgment and patient report during the visit. The study also did not include standardized neurologic examination, nerve conduction studies, quantitative sensory testing, or tissue-level validation. This limits the ability to distinguish patient-reported symptom burden, clinician-recorded toxicity grade, and objective neurologic impairment. Fifth, dose modifications were not available in a fully adjudicated, subject-level, time-varying dataset, limiting formal analysis of delivered vincristine exposure, dose reduction, and dose omission as dynamic clinical variables. In addition, dose changes can occur for reasons other than development of CIPN, so such modifications cannot be assumed to be neuropathy-related in every case; they are best interpreted as real-world treatment modifications that may alter subsequent vincristine exposure, NfL release, and symptom trajectories. Finally, T5/Post data were exploratory, PRO-based, and available only in a small subset; they should not be used to estimate the prevalence of chronic CIPN or to assess predictive performance.

Despite these limitations, this study provides an important prospective reference point for interpreting plasma NfL in adults receiving vincristine-containing lymphoma therapy. The cohort captures a clinically realistic treatment setting, with pre-cycle sampling, patient-reported symptom assessment, routine-care CTCAE toxicity grading, BPI severity assessment, dose-modification context, and exploratory long-term PRO follow-up. The robust early increase in plasma NfL supports its sensitivity to vincristine-containing treatment exposure and neuroaxonal injury biology, while the incomplete alignment among NfL, CIPN20, CTCAE toxicity, BPI severity, and dose modification demonstrates why NfL should not be interpreted as a stand-alone symptom or toxicity measure. In this respect, the study is valuable not because it establishes a predictive biomarker, but because it defines the key methodological and interpretive issues that larger vincristine-focused CIPN biomarker studies must address. Future studies should use standardized pretreatment sampling, larger longitudinal cohorts, post-treatment and survivorship follow-up, prespecified lagged analyses, subject-level dose-intensity data, and clinician-confirmed neurologic endpoints. Larger cohort studies incorporating clinical assessments are needed before NfL can be used to inform routine CIPN clinical decisions [16].

## Conclusion

Plasma NfL increased substantially during first-line vincristine-containing CHOP- or EPOCH-based chemotherapy in adults with NHL, with an early rise and sustained elevation through later chemotherapy exposure. Patient-reported neuropathy symptom burden also increased during treatment. However, NfL did not serve as a direct surrogate for patient-reported symptom severity, clinician-recorded toxicity, pain severity, or dose-modification context at any single timepoint. Exploratory analysis revealed a possible association between early NfL changes and later CIPN20 worsening. Larger studies are needed to determine whether longitudinal NfL trajectories can help predict CIPN symptom burden and inform intervention strategies for adults with NHL.

## Data Availability

All data produced in the present work are contained in the manuscript.
Additional relevant data or analysis in the present study are available upon request to the authors.

## Acknowledgments

We are grateful to the patients who participated in this study. We thank the Hematology Tissue Bank Shared Resource and the Recruitment, Intervention and Survey Shared Resource at The Ohio State University Comprehensive Cancer Center for research blood processing, REDcap database development and study support. Finally, we would like to thank Dr. Bhuvana Ramaswamy for her invaluable support.

## Ethics approval

This study was performed in accordance with the principles of the Declaration of Helsinki. The study protocol was approved by The Ohio State University Institutional Review Board.

## Consent to participate

Written informed consent was obtained from all individual participants included in the study.

## Consent for publication

Not applicable. The manuscript does not contain identifying individual participant information or identifiable images. Participant codes shown in figures and supplementary tables are randomized public analysis codes and do not correspond to study IDs or clinical identifiers.

## Data availability

Deidentified data supporting the findings of this study are included in the manuscript and supplementary materials. Additional deidentified data may be available from the corresponding author upon reasonable request and subject to institutional and IRB requirements.

## Code availability

Analysis code may be available from the corresponding author upon reasonable request and subject to institutional and IRB requirements.

## Clinical trial registration

Not applicable. This was an observational cohort study and participants were not assigned to an intervention.

## Author contributions

G.A.M. designed the study, coordinated the work, interpreted the data, wrote the initial manuscript, and revised the manuscript. G.J.S. contributed to conceptual framing, interpretation of results, statistical analyses, and writing and revision of the manuscript. L.W.-C. contributed to conceptual framing, statistical interpretation, and writing and revision of the manuscript. P.M.S. performed and advised on statistical analyses. L.F. and S.S.K. contributed to data collection. T.V., R.B., D.B., B.C., K.M., and Y.S. contributed to patient care and clinical interpretation. M.B.L. contributed to study conception, clinical interpretation, and manuscript review. All authors read and approved the final manuscript.

## Supplemental figure and table legends

**Figure S1.**
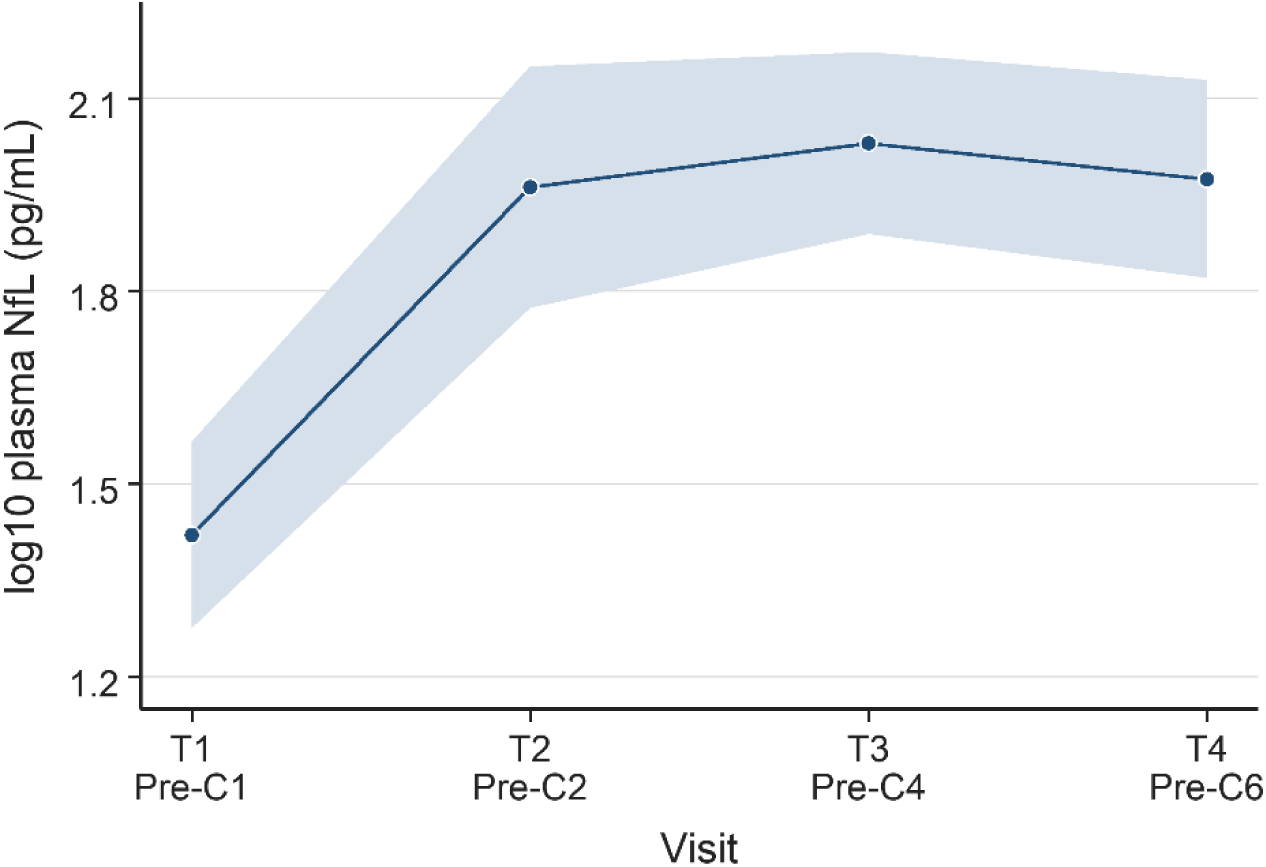
Primary NfL model displayed on the log10 scale. Model-estimated plasma NfL and 95% confidence intervals from the primary mixed-effects model shown directly on the log10 scale. The model included timepoint as a fixed effect and a participant-level random intercept. Points show model estimates and the shaded band shows the 95% confidence interval. P values are model-based estimated marginal mean contrast p values comparing each follow-up timepoint with T1/Pre-C1. Compared with T1/Pre-C1, log10 NfL was higher at T2/Pre-C2 (p = 4.16e-06) T3/Pre-C4 (p = 6.41e-10) and T4/Pre-C6 (p = 3.39e-08).

**Figure S2.**
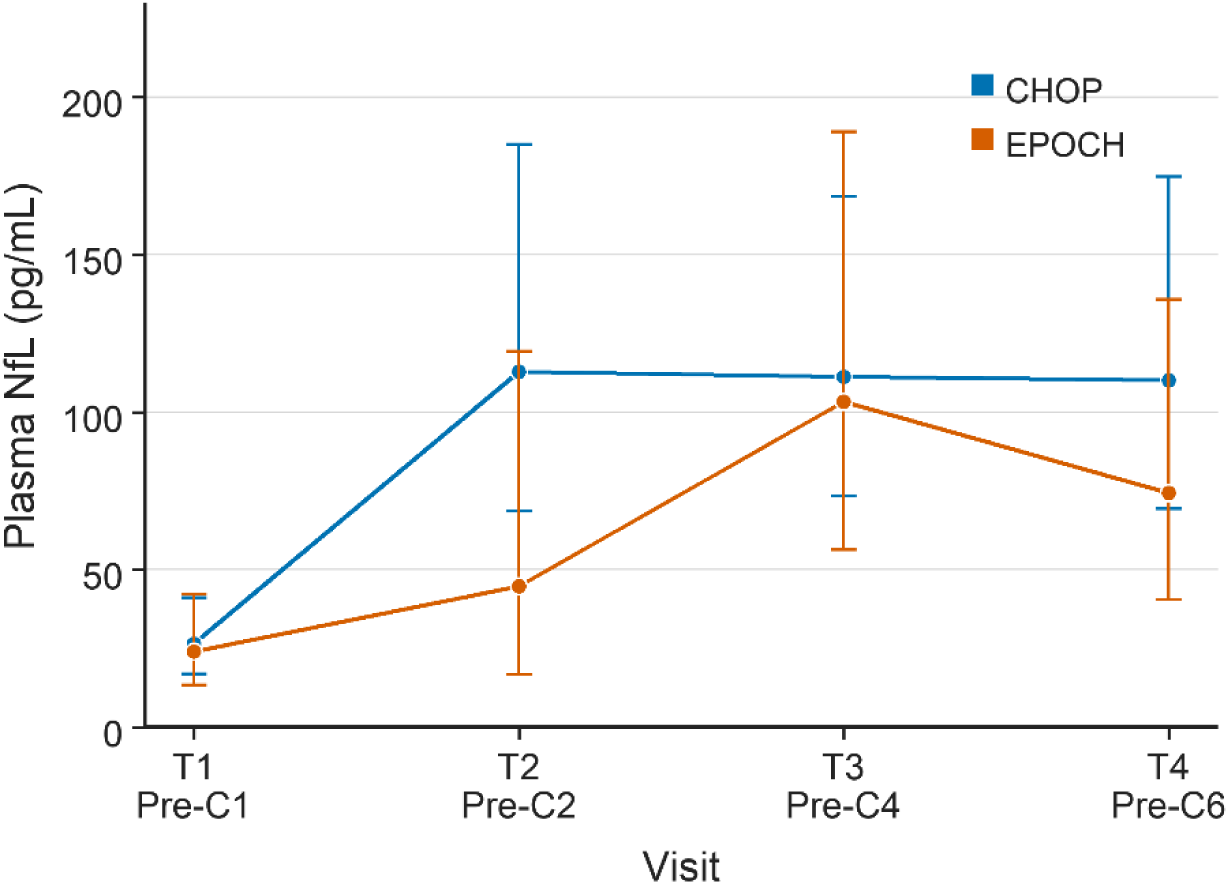
Plasma NfL trajectories by chemotherapy regimen. Model-estimated plasma NfL geometric means and 95% confidence intervals by chemotherapy regimen, back-transformed to pg/mL from an exploratory mixed-effects model including timepoint, regimen, and timepoint-by-regimen interaction terms. Points show back-transformed model estimates, and vertical error bars show 95% confidence intervals. Statistical testing used model-based fixed-effect and interaction terms from the exploratory regimen model. The main effect for EPOCH compared with CHOP was not significant (p = 0.779). Timepoint-by-regimen interaction terms were not significant at T2/Pre-C2 (p = 0.132) T3/Pre-C4 (p = 0.929) or T4/Pre-C6 (p = 0.356) compared with T1/Pre-C1. CHOP, cyclophosphamide, doxorubicin, vincristine, and prednisone; EPOCH, etoposide, prednisone, vincristine, cyclophosphamide, and doxorubicin; NfL, neurofilament light chain.

**Figure S3.**
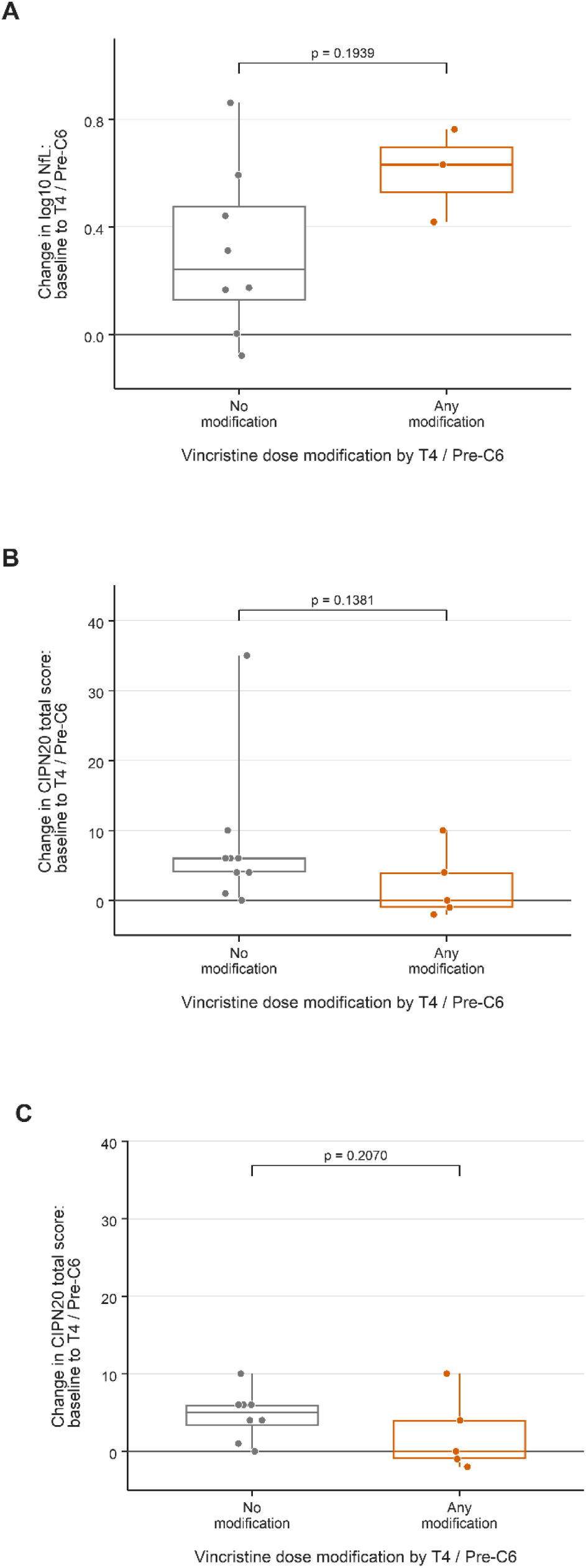
Exploratory vincristine dose-modification analyses. (A) Change in log10 plasma NfL from baseline to T4/Pre-C6 stratified by whether participants had any vincristine dose modification by T4/Pre-C6. (B) Change in raw CIPN20 total score from baseline to T4/Pre-C6 stratified by any vincristine dose modification (including discontinuation) by T4/Pre-C6. Boxes show the interquartile range, horizontal lines show the median, whiskers show the observed minimum and maximum, and points show individual participants. Mann-Whitney U tests. (A) all-participant log10 NfL analysis included n = 11 (3 dose-modified, 8 with no modification) p = 0.1939. (B) all-participant CIPN20 analysis included n = 14 (5 dose-modified, 9 with no modification) p = 0.1381. The high CIPN20-change outlier was in the no-modification group with delta CIPN20 = +35 from baseline to T4/Pre-C6; this was an IQR-rule outlier within the no-modification group but was retained because it may associate with an observed clinical value. NfL, neurofilament light chain; CIPN20, Chemotherapy-Induced Peripheral Neuropathy questionnaire.ΔΔ

**Table S1.** Timepoint-specific associations between plasma NfL and CIPN20 scores. Spearman correlations between paired log10 plasma NfL and CIPN20 total score are shown separately forT1/Pre-C1,T2/Pre-C2,T3/Pre-C4, andT4/Pre-C6. These analyses were exploratory and underpowered, and were used to contextualize the pooled visit-level association shown in Figure 5A. NfL, neurofilament light chain; CIPN20, Chemotherapy-Induced Peripheral Neuropathy questionnaire.

| Timepoint | N | Spearman $\rho$ | p value |
| --- | --- | --- | --- |
| T1/Pre-C1 | 16 | -0.358 | 0.174 |
| T2/Pre-C2 | 7 | 0.891 | 0.007 |
| T3/Pre-C4 | 17 | 0.399 | 0.112 |
| T4/Pre-C6 | 11 | -0.119 | 0.728 |

**Table S2.** Subject-level vincristine dose-modification and treatment-period clinical context. Rows show subject-level visit data for vincristine dose modification, CIPN20 total and sensory scores, BPI-SF, CTCAE, NfL, and the added baseline cancer stage and HbA1c/eA1c context. HbA1c/eA1c combines measured HbA1c with glucose-derived estimated HbA1c when HbA1c was unavailable; glucose-derived eA1c values were used descriptively only. ND: not documented; CIPN20, Chemotherapy-Induced Peripheral Neuropathy 20-item questionnaire; BPI-SF, Brief Pain Inventory-Short Form; CTCAE, Common Terminology Criteria for Adverse Events; NfL, neurofilament light chain; HbA1c, hemoglobin A1c; eA1c, estimated HbA1c.

| Code | Treatment | Stage | HbA1c/<br>eA1c<br>(%) | HbA1c/eA1c<br>source | HbA1c/eA1c<br>category | Timepoint | Vincristine<br>dose<br>modification | Dose-modification<br>reason | CIPN<br>20<br>total | CIPN<br>20<br>sensory | BPI-SF | CTCAE | NfL<br>(pg/mL) |
| --- | --- | --- | --- | --- | --- | --- | --- | --- | --- | --- | --- | --- | --- |
| P01 | CHOP | IV | 5.6 | Measured<br>HbA1c | Normal | T1/Pre-C1 | No modification | Not applicable | 19 | 9 | 1.5 | 0 | 69.9 |
| P02 | CHOP | IV | 6.9 | Glucose-derived eA1c | Diabetes<br>range | T1/Pre-C1 | No modification | Not applicable | 21 | 10 | 1 | 0 | 187.9 |
| P03 | CHOP | IV | 5.1 | Measured<br>HbA1c | Normal | T2/Pre-C2 | No modification | Not applicable | 25 | 10 | 2.5 | 0 | 101.6 |
| P03 | CHOP | IV | 5.1 | Measured<br>HbA1c | Normal | T3/Pre-C4 | No modification | Not applicable | 25 | 11 | 4.75 | 0 | 96.3 |
| P03 | CHOP | IV | 5.1 | Measured<br>HbA1c | Normal | T4/Pre-C6 | No modification | Not applicable | 31 | 14 | 5.25 | 0 | Missing |
| P04 | EPOCH | IV | 5.4 | Measured<br>HbA1c | Normal | T1/Pre-C1 | No modification | Not applicable | 25 | 10 | 0 | 1 | 12.9 |
| P04 | EPOCH | IV | 5.4 | Measured<br>HbA1c | Normal | T3/Pre-C4 | Removed | CIPN | 32 | 14 | 0 | 2 | 175.5 |
| P04 | EPOCH | IV | 5.4 | Measured<br>HbA1c | Normal | T4/Pre-C6 | Removed | CIPN | 35 | 13 | 0 | 1 | 75 |
| P05 | CHOP | IV | 7.0 | Measured<br>HbA1c | Diabetes<br>range | T1/Pre-C1 | No modification | Not applicable | 19 | 9 | 0 | 0 | 17.9 |
| P05 | CHOP | IV | 7.0 | Measured<br>HbA1c | Diabetes<br>range | T3/Pre-C4 | No modification | Not applicable | 27 | 9 | 1.75 | 0 | 70.3 |
| P06 | EPOCH | II | 4.7 | Glucose-derived eA1c | Normal | T1/Pre-C1 | No modification | Not applicable | 21 | 9 | 0 | 0 | 63.1 |
| P06 | EPOCH | II | 4.7 | Glucose-derived eA1c | Normal | T3/Pre-C4 | No modification | Not applicable | 19 | 9 | 0 | 0 | Missing |
| P06 | EPOCH | II | 4.7 | Glucose-derived eA1c | Normal | T4/Pre-C6 | No modification | Not applicable | 25 | 10 | 0 | 0 | 94.4 |
| P07 | CHOP | III | 6.0 | Measured<br>HbA1c | Prediabetes | T1/Pre-C1 | No modification | Not applicable | 19 | 9 | 0 | 0 | 23.7 |
| P07 | CHOP | III | 6.0 | Measured<br>HbA1c | Prediabetes | T3/Pre-C4 | No modification | Not applicable | 20 | 9 | 2 | 0 | 114.3 |
| P07 | CHOP | III | 6.0 | Measured<br>HbA1c | Prediabetes | T4/Pre-C6 | No modification | Not applicable | 25 | 10 | 0.75 | 1 | 172.1 |
| P08 | EPOCH | ND | 5.4 | Measured<br>HbA1c | Normal | T1/Pre-C1 | No modification | Not applicable | 25 | 10 | Miss<br>ing | 0 | Missing |
| P09 | EPOCH | ND | 4.2 | Measured<br>HbA1c | Normal | T1/Pre-C1 | No modification | Not applicable | 20 | 10 | 0.25 | 0 | 72.4 |
| P09 | EPOCH | ND | 4.2 | Measured<br>HbA1c | Normal | T3/Pre-C4 | No modification | Not applicable | 21 | 10 | 0.5 | 1 | Missing |
| P09 | EPOCH | ND | 4.2 | Measured<br>HbA1c | Normal | T4/Pre-C6 | 50% reduction | constipation | 20 | 10 | 0.25 | 1 | 189.9 |
| P10 | CHOP | IV | 7.3 | Measured<br>HbA1c | Diabetes<br>range | T2/Pre-C2 | No modification | Not applicable | 21 | 10 | 0 | 0 | 80.1 |
| P10 | CHOP | IV | 7.3 | Measured<br>HbA1c | Diabetes<br>range | T3/Pre-C4 | No modification | Not applicable | 25 | 11 | 0 | 0 | 80 |
| P10 | CHOP | IV | 7.3 | Measured<br>HbA1c | Diabetes<br>range | T4/Pre-C6 | No modification | Not applicable | 27 | 12 | 0 | 0 | 80.8 |
| P11 | CHOP | IV | 6.8 | Measured<br>HbA1c | Diabetes<br>range | T1/Pre-C1 | No modification | Not applicable | 20 | 9 | 0 | Missing | 31.4 |
| P11 | CHOP | IV | 6.8 | Measured<br>HbA1c | Diabetes<br>range | T3/Pre-C4 | No modification | Not applicable | 21 | 9 | 0 | 0 | 232.2 |
| P12 | EPOCH | ND | 6.0 | Measured<br>HbA1c | Prediabetes | T1/Pre-C1 | No modification | Not applicable | 22 | 11 | 0 | 0 | 58.9 |
| P13 | CHOP | ND | 6.5 | Glucose-derived eA1c | Diabetes<br>range | T1/Pre-C1 | No modification | Not applicable | 21 | 9 | 2.5 | 0 | 16.1 |
| P13 | CHOP | ND | 6.5 | Glucose-derived eA1c | Diabetes<br>range | T3/Pre-C4 | No modification | Not applicable | 26 | 15 | 0 | 1 | 99.9 |
| P13 | CHOP | ND | 6.5 | Glucose-derived eA1c | Diabetes<br>range | T4/Pre-C6 | No modification | Not applicable | 31 | 14 | 0 | 1 | 63.2 |
| P14 | CHOP | IV | 6.0 | Measured<br>HbA1c | Prediabetes | T2/Pre-C2 | 25% reduction | CIPN | 21 | 10 | 0.5 | 2 | 74.3 |
| P14 | CHOP | IV | 6.0 | Measured<br>HbA1c | Prediabetes | T3/Pre-C4 | 25% reduction | CIPN | 23 | 11 | 4 | 1 | Missing |
| P14 | CHOP | IV | 6.0 | Measured<br>HbA1c | Prediabetes | T4/Pre-C6 | 50% reduction | CIPN | 25 | 12 | 4.5 | 2 | Missing |
| P15 | CHOP | IV | 4.7 | Glucose-derived eA1c | Normal | T2/Pre-C2 | No modification | Not applicable | 19 | 9 | 0 | 0 | 68.7 |
| P15 | CHOP | IV | 4.7 | Glucose-derived eA1c | Normal | T3/Pre-C4 | No modification | Not applicable | 18 | 9 | 0 | 0 | 62.6 |
| P15 | CHOP | IV | 4.7 | Glucose-derived eA1c | Normal | T4/Pre-C6 | No modification | Not applicable | 19 | 9 | 0 | 0 | 57.4 |
| P16 | CHOP | IIIA | 6.2 | Measured<br>HbA1c | Prediabetes | T1/Pre-C1 | No modification | Not applicable | 20 | 9 | 0 | Missing | 19.7 |
| P16 | CHOP | IIIA | 6.2 | Measured<br>HbA1c | Prediabetes | T3/Pre-C4 | 50% reduction | Multifactorial | 24 | 9 | 1.75 | 1 | 74.8 |
| P16 | CHOP | IIIA | 6.2 | Measured<br>HbA1c | Prediabetes | T4/Pre-C6 | 50% reduction | Multifactorial | 19 | 9 | 4 | 0 | 84.3 |
| P17 | EPOCH | IV | 4.3 | Measured<br>HbA1c | Normal | T1/Pre-C1 | No modification | Not applicable | 20 | 9 | 0 | 0 | 19.2 |
| P17 | EPOCH | IV | 4.3 | Measured<br>HbA1c | Normal | T3/Pre-C4 | No modification | Not applicable | 20 | 9 | 0 | 0 | 58 |
| P17 | EPOCH | IV | 4.3 | Measured<br>HbA1c | Normal | T4/Pre-C6 | No modification | Not applicable | 21 | 9 | 0 | 0 | 53.1 |
| P18 | EPOCH | IV | 4.5 | Measured<br>HbA1c | Normal | T2/Pre-C2 | No modification | Not applicable | 19 | 9 | 0 | 0 | 36.2 |
| P18 | EPOCH | IV | 4.5 | Measured<br>HbA1c | Normal | T3/Pre-C4 | No modification | Not applicable | 35 | 20 | 7.75 | 1 | 63.9 |
| P18 | EPOCH | IV | 4.5 | Measured<br>HbA1c | Normal | T4/Pre-C6 | No modification | Not applicable | 54 | 24 | 4 | 0 | 74.4 |
| P19 | CHOP | IV | 6.3 | Glucose-derived eA1c | Prediabetes | T2/Pre-C2 | No modification | Not applicable | 20 | 9 | Miss<br>ing | 0 | 57.7 |
| P19 | CHOP | IV | 6.3 | Glucose-derived eA1c | Prediabetes | T3/Pre-C4 | No modification | Not applicable | 21 | 9 | 0 | 0 | Missing |
| P19 | CHOP | IV | 6.3 | Glucose-derived eA1c | Prediabetes | T4/Pre-C6 | No modification | Not applicable | 24 | 9 | 0 | 0 | 84.6 |
| P20 | EPOCH | ND | 5.5 | Glucose-derived eA1c | Normal | T1/Pre-C1 | No modification | Not applicable | 22 | 9 | 0 | 0 | 12.1 |
| P20 | EPOCH | ND | 5.5 | Glucose-derived eA1c | Normal | T3/Pre-C4 | Removed | Ileus | 18 | 9 | 0 | 0 | 32.1 |
| P20 | EPOCH | ND | 5.5 | Glucose-derived eA1c | Normal | T4/Pre-C6 | Removed | Ileus | 20 | 9 | 0 | 0 | Missing |

| Code | Treatment | Stage | HbA1c/eA1c (%) | HbA1c/eA1c source | HbA1c/eA1c category | Timepoint | Vincristine dose modification | Dose-modification reason | CIPN 20 total | CIPN 20 sensory | BPI-SF | CTCAE | NfL (pg/mL) |
| --- | --- | --- | --- | --- | --- | --- | --- | --- | --- | --- | --- | --- | --- |
| P21 | CHOP | IV | 8.5 | Measured HbA1c | Diabetes range | T2/Pre-C2 | No modification | Not applicable | 22 | 9 | 0 | 0 | 142.2 |
| P21 | CHOP | IV | 8.5 | Measured HbA1c | Diabetes range | T3/Pre-C4 | No modification | Not applicable | 21 | 9 | 0 | 1 | 67.4 |
| P22 | CHOP | ND | 4.7 | Measured HbA1c | Normal | T2/Pre-C2 | No modification | Not applicable | 22 | 11 | 0 | 0 | Missing |
| P22 | CHOP | ND | 4.7 | Measured HbA1c | Normal | T3/Pre-C4 | No modification | Not applicable | 20 | 9 | 0 | 0 | 37 |
| P22 | CHOP | ND | 4.7 | Measured HbA1c | Normal | T4/Pre-C6 | No modification | Not applicable | Missing | Missing | Missing | 0 | 34 |
| P23 | EPOCH | IIA | 5.4 | Measured HbA1c | Normal | T1/Pre-C1 | No modification | Not applicable | 21 | 9 | 0 | 0 | 6.8 |
| P23 | EPOCH | IIA | 5.4 | Measured HbA1c | Normal | T3/Pre-C4 | No modification | Not applicable | 26 | 13 | 0 | 0 | 65.7 |
| P24 | CHOP | III-IV | 5.4 | Measured HbA1c | Normal | T1/Pre-C1 | 25% reduction | Not specified | 18 | 9 | Missing | 0 | 71.5 |
| P24 | CHOP | III-IV | 5.4 | Measured HbA1c | Normal | T3/Pre-C4 | 25% reduction | Not specified | 26 | 10 | 2.25 | 0 | 212.8 |
| P25 | CHOP | ND | 5.0 | Measured HbA1c | Normal | T1/Pre-C1 | No modification | Not applicable | 21 | 9 | 3.25 | 0 | 25.9 |
| P25 | CHOP | ND | 5.0 | Measured HbA1c | Normal | T3/Pre-C4 | No modification | Not applicable | 25 | 9 | 5 | 0 | 208.6 |

**Table S3.** Exploratory T5/Post patient-reported outcomes and dose-modification context. Long-term patient-reported outcomes are shown for participants with available T5/Post data 24-42 months after completion of vincristine-containing chemotherapy. Treatment-period CIPN20, T5/Post CIPN20, T5/Post BPI-SF, T3 CTCAE grade, available treatment-period plasma NfL values, and vincristine dose-modification history are presented descriptively. NfL, neurofilament light chain; CIPN20, Chemotherapy-Induced Peripheral Neuropathy questionnaire; CTCAE, Common Terminology Criteria for Adverse Events; BPI-SF, Brief Pain Inventory-Short Form.

| Code | Treatment | Months since completion | T4/Pre-C6 CIPN20 total | T5/Post CIPN20 total | T5/Post CIPN20 sensory | T5/Post BPI-SF | T4/Pre-C6 CTCAE | Dose modification | % reduction/removal | Timing/reason | NfL T1/Pre-C1 | NfL T2/Pre-C2 | NfL T3/Pre-C4 | NfL T4/Pre-C6 |
| --- | --- | --- | --- | --- | --- | --- | --- | --- | --- | --- | --- | --- | --- | --- |
| P06 | EPOCH | 28 | 25 | 20 | 9 | 0.25 | 0 | No | No modification |  | 63.1 |  |  | 94.4 |
| P09 | EPOCH | 42 | 20 | 28 | 16 | 0 | 1 | Yes | 50% reduction | C6; constipation | 72.4 |  |  | 189.9 |
| P13 | CHOP | 38 | 31 | 25 | 13 | 3.5 | 1 | No | No modification |  | 16.1 |  | 99.9 | 63.2 |
| P14 | CHOP | 24 | 25 | 36 | 21 | 4.25 | 2 | Yes | 25% reduction, 50% reduction | C2/C5; CIPN |  | 74.3 |  |  |
| P16 | CHOP | 39 | 19 | 21 | 10 | 1.5 | 0 | Yes | 50% reduction | C4; multifactorial | 19.7 |  | 74.8 | 84.3 |
| P18 | EPOCH | 38 | 54 | 22 | 9 | 2.5 | 0 | No | No modification |  |  | 36.2 | 63.9 | 74.4 |

## References

1. Bartlett NL, Wilson WH, Jung S-H, et al (2019) Dose-Adjusted EPOCH-R Compared With R-CHOP as Frontline Therapy for Diffuse Large B-Cell Lymphoma: Clinical Outcomes of the Phase III Intergroup Trial Alliance/CALGB 50303. J Clin Oncol 37:1790–1799. 10.1200/JCO.18.01994

2. Madsen ML, Due H, Ejskjær N, et al (2019) Aspects of vincristine-induced neuropathy in hematologic malignancies: a systematic review. Cancer Chemother Pharmacol 84:471–485. 10.1007/s00280-019-03884-5

3. Li T, Trinh T, Bosco A, et al (2024) Characterising vincristine-induced peripheral neuropathy in adults: symptom development and long-term persistent outcomes. Support Care Cancer 32:278. 10.1007/s00520-024-08484-5

4. Okada N, Hanafusa T, Sakurada T, et al (2014) Risk Factors for Early-Onset Peripheral Neuropathy Caused by Vincristine in Patients With a First Administration of R-CHOP or R-CHOP-Like Chemotherapy. J Clin Med Res. 10.14740/jocmr1856w

5. Sawaki A, Miyazaki K, Yamaguchi M, et al (2020) Genetic polymorphisms and vincristine-induced peripheral neuropathy in patients treated with rituximab, cyclophosphamide, doxorubicin, vincristine, and prednisone therapy. Int J Hematol 111:686–691. 10.1007/s12185-020-02832-x

6. Su Y-C, Lai Y-H, Hsieh S-T, et al (2024) Acute, long-term or non-vincristine-induced peripheral neuropathy among non-Hodgkin lymphoma survivors: Symptoms, daily activities, functional status, and quality of life. European Journal of Oncology Nursing 69:102540. 10.1016/j.ejon.2024.102540

7. Kim B-J, Park H-R, Roh HJ, et al (2010) Chemotherapy-related polyneuropathy may deteriorate quality of life in patients with B-cell lymphoma. Qual Life Res 19:1097–1103. 10.1007/s11136-010-9670-0

8. Loprinzi CL, Lacchetti C, Bleeker J, et al (2020) Prevention and Management of Chemotherapy-Induced Peripheral Neuropathy in Survivors of Adult Cancers: ASCO Guideline Update. J Clin Oncol 38:3325–3348. 10.1200/JCO.20.01399

9. Hatzl S, Posch F, Rezai A, et al (2021) Vinorelbine as substitute for vincristine in patients with diffuse large B cell lymphoma and vincristine-induced neuropathy. Support Care Cancer 29:5197–5207. 10.1007/s00520-021-06059-2

10. LaPointe NE, Morfini G, Brady ST, et al (2013) Effects of eribulin, vincristine, paclitaxel and ixabepilone on fast axonal transport and kinesin-1 driven microtubule gliding: implications for chemotherapy-induced peripheral neuropathy. Neurotoxicology 37:231–239. 10.1016/j.neuro.2013.05.008

11. Jordan B, Margulies A, Cardoso F, et al (2020) Systemic anticancer therapy-induced peripheral and central neurotoxicity: ESMO-EONS-EANO Clinical Practice Guidelines for diagnosis, prevention, treatment and follow-up. Ann Oncol 31:1306–1319. 10.1016/j.annonc.2020.07.003

12. Jordan MA, Thrower D, Wilson L (1991) Mechanism of inhibition of cell proliferation by Vinca alkaloids. Cancer Res 51:2212–2222

13. Khalil M, Teunissen CE, Otto M, et al (2018) Neurofilaments as biomarkers in neurological disorders. Nat Rev Neurol 14:577–589. 10.1038/s41582-018-0058-z

14. Yuan A, Rao MV, Veeranna, Nixon RA (2017) Neurofilaments and Neurofilament Proteins in Health and Disease. Cold Spring Harb Perspect Biol 9:a018309. 10.1101/cshperspect.a018309

15. Andersen NE, Boehmerle W, Huehnchen P, Stage TB (2024) Neurofilament light chain as a biomarker of chemotherapy-induced peripheral neuropathy. Trends Pharmacol Sci 45:872–879. 10.1016/j.tips.2024.08.001

16. Alberti P, Faithfull S, Argyriou AA, et al (2026) Serum neurofilament light chain (sNFL) as a predictive biomarker for Chemotherapy-induced peripheral neurotoxicity (CIPN): consideration of current evidence, validation steps, and barriers to CIPN biomarker implementation in clinical practice—a narrative review. Support Care Cancer 34:508. 10.1007/s00520-026-10690-2

17. Cavaletti G, Pizzamiglio C, Man A, et al (2023) Studies to Assess the Utility of Serum Neurofilament Light Chain as a Biomarker in Chemotherapy-Induced Peripheral Neuropathy. Cancers (Basel) 15:4216. 10.3390/cancers15174216

18. Huehnchen P, Schinke C, Bangemann N, et al (2022) Neurofilament proteins as a potential biomarker in chemotherapy-induced polyneuropathy. JCI Insight 7:e154395. 10.1172/jci.insight.154395

19. Burgess BL, Cho E, Honigberg L (2022) Neurofilament light as a predictive biomarker of unresolved chemotherapy-induced peripheral neuropathy in subjects receiving paclitaxel and carboplatin. Sci Rep 12:15593. 10.1038/s41598-022-18716-5

20. Velasco R, Marco C, Domingo-Domenech E, et al (2024) Plasma neurofilament light chain levels in chemotherapy-induced peripheral neurotoxicity according to type of anticancer drug. Euro J of Neurology 31:e16369. 10.1111/ene.16369

21. Kim S-H, Choi MK, Park NY, et al (2020) Serum neurofilament light chain levels as a biomarker of neuroaxonal injury and severity of oxaliplatin-induced peripheral neuropathy. Sci Rep 10:7995. 10.1038/s41598-020-64511-5

22. Meregalli C, Fumagalli G, Alberti P, et al (2020) Neurofilament light chain: a specific serum biomarker of axonal damage severity in rat models of Chemotherapy-Induced Peripheral Neurotoxicity. Arch Toxicol 94:2517–2522. 10.1007/s00204-020-02755-w

23. Stephan LU, Abel J, Boehmerle W, et al (2025) Chemotherapy-induced polyneuropathy: diagnostic challenges and the potential of neurofilament as a biomarker for sensory disorders: the CONKO 023-ChemTox Trial. J Neurol 272:719. 10.1007/s00415-025-13463-9

24. Akamine S, Marutani N, Kanayama D, et al (2020) Renal function is associated with blood neurofilament light chain level in older adults. Sci Rep 10:20350. 10.1038/s41598-020-76990-7

25. Bavato F, Barro C, Schnider LK, et al (2024) Introducing neurofilament light chain measure in psychiatry: current evidence, opportunities, and pitfalls. Mol Psychiatry 29:2543–2559. 10.1038/s41380-024-02524-6

26. Polymeris AA, Helfenstein F, Benkert P, et al (2022) Renal Function and Body Mass Index Contribute to Serum Neurofilament Light Chain Levels in Elderly Patients With Atrial Fibrillation. Front Neurosci 16:819010. 10.3389/fnins.2022.819010

27. Simrén J, Andreasson U, Gobom J, et al (2022) Establishment of reference values for plasma neurofilament light based on healthy individuals aged 5-90 years. Brain Commun 4:fcac174. 10.1093/braincomms/fcac174

28. Harris PA, Taylor R, Thielke R, et al (2009) Research electronic data capture (REDCap)—A metadata-driven methodology and workflow process for providing translational research informatics support. Journal of Biomedical Informatics 42:377–381. 10.1016/j.jbi.2008.08.010

29. Harris PA, Taylor R, Minor BL, et al (2019) The REDCap consortium: Building an international community of software platform partners. Journal of Biomedical Informatics 95:103208. 10.1016/j.jbi.2019.103208

30. Postma TJ, Aaronson NK, Heimans JJ, et al (2005) The development of an EORTC quality of life questionnaire to assess chemotherapy-induced peripheral neuropathy: The QLQ-CIPN20. European Journal of Cancer 41:1135–1139. 10.1016/j.ejca.2005.02.012

31. Lavoie Smith EM, Barton DL, Qin R, et al (2013) Assessing patient-reported peripheral neuropathy: the reliability and validity of the European Organization for Research and Treatment of Cancer QLQ-CIPN20 Questionnaire. Qual Life Res 22:2787–2799. 10.1007/s11136-013-0379-8

32. Kieffer JM, Postma TJ, Van De Poll-Franse L, et al (2017) Evaluation of the psychometric properties of the EORTC chemotherapy-induced peripheral neuropathy questionnaire (QLQ-CIPN20). Qual Life Res 26:2999–3010. 10.1007/s11136-017-1626-1

33. Cleeland CS, Ryan KM (1994) Pain assessment: global use of the Brief Pain Inventory. Ann Acad Med Singap 23:129–138

34. Dworkin RH, Turk DC, Farrar JT, et al (2005) Core outcome measures for chronic pain clinical trials: IMMPACT recommendations. Pain 113:9–19. 10.1016/j.pain.2004.09.012

35. National Cancer Institute (2017) Common Terminology Criteria for Adverse Events (CTCAE), Version 5.0. National Cancer Institute

36. Nathan DM, Kuenen J, Borg R, et al (2008) Translating the A1C Assay Into Estimated Average Glucose Values. Diabetes Care 31:1473–1478. 10.2337/dc08-0545

37. Barker HL, Morrison D, Llano A, et al (2023) Practical Guide to Glucocorticoid Induced Hyperglycaemia and Diabetes. Diabetes Ther 14:937–945. 10.1007/s13300-023-01393-6

38. American Diabetes Association Professional Practice Committee, ElSayed NA, Aleppo G, et al (2024) 6. Glycemic Goals and Hypoglycemia: *Standards of Care in Diabetes—*2024. Diabetes Care 47:S111–S125. 10.2337/dc24-S006

39. Worthen-Chaudhari LC, Crasta JE, Schnell PM, et al (2025) Neurologic dance training and home exercise improve motor-cognitive dual-task function similarly, but through potentially different mechanisms, among breast cancer survivors with chemotherapy-induced neuropathy: Initial results of a randomized, controlled clinical trial. J Alzheimers Dis 105:1114–1130. 10.1177/13872877241291440

40. Moriyama B, Henning SA, Leung J, et al (2012) Adverse interactions between antifungal azoles and vincristine: review and analysis of cases. Mycoses 55:290–297. 10.1111/j.1439-0507.2011.02158.x

41. Dennison JB, Kulanthaivel P, Barbuch RJ, et al (2006) Selective metabolism of vincristine in vitro by CYP3A5. Drug Metabolism and Disposition 34:1317–1327. 10.1124/dmd.106.009902

42. McNally GA, Aossey CM, Wiczer T, et al (2024) A retrospective cohort study describing chemotherapy-induced peripheral neuropathy in Non-Hodgkin lymphoma patients treated with EPOCH ± R: does HIV status matter? Leukemia & Lymphoma 65:1110–1116. 10.1080/10428194.2024.2340051

43. Kahn OI, Dominguez SL, Glock C, et al (2025) Secreted neurofilament light chain after neuronal damage induces myeloid cell activation and neuroinflammation. Cell Reports 44:115382. 10.1016/j.celrep.2025.115382

44. Kiguchi N, Maeda T, Kobayashi Y, et al (2008) The critical role of invading peripheral macrophage-derived interleukin-6 in vincristine-induced mechanical allodynia in mice. European Journal of Pharmacology 592:87–92. 10.1016/j.ejphar.2008.07.008

45. Ji X-T, Qian N-S, Zhang T, et al (2013) Spinal Astrocytic Activation Contributes to Mechanical Allodynia in a Rat Chemotherapy-Induced Neuropathic Pain Model. PLoS ONE 8:e60733. 10.1371/journal.pone.0060733

46. Montague K, Simeoli R, Valente J, Malcangio M (2018) A novel interaction between CX3CR1 and CCR2 signalling in monocytes constitutes an underlying mechanism for persistent vincristine-induced pain. J Neuroinflammation 15:101. 10.1186/s12974-018-1116-6

47. Old EA, Nadkarni S, Grist J, et al (2014) Monocytes expressing CX3CR1 orchestrate the development of vincristine-induced pain. J Clin Invest 124:2023–2036. 10.1172/JCI71389

48. Starobova H, Mueller A, Allavena R, et al (2019) Minocycline Prevents the Development of Mechanical Allodynia in Mouse Models of Vincristine-Induced Peripheral Neuropathy. Front Neurosci 13:653. 10.3389/fnins.2019.00653

49. Starobova H, Monteleone M, Adolphe C, et al (2021) Vincristine-induced peripheral neuropathy is driven by canonical NLRP3 activation and IL-1β release. J Exp Med 218:e20201452. 10.1084/jem.20201452

50. Le-Rademacher J, Kanwar R, Seisler D, et al (2017) Patient-reported (EORTC QLQ-CIPN20) versus physician-reported (CTCAE) quantification of oxaliplatin- and paclitaxel/carboplatin-induced peripheral neuropathy in NCCTG/Alliance clinical trials. Support Care Cancer 25:3537–3544. 10.1007/s00520-017-3780-y

51. Tan AC, McCrary JM, Park SB, et al (2019) Chemotherapy-induced peripheral neuropathy—patient-reported outcomes compared with NCI-CTCAE grade. Support Care Cancer 27:4771–4777. 10.1007/s00520-019-04781-6

52. Alberti P, Bernasconi DP, Cornblath DR, et al (2021) Prospective Evaluation of Health Care Provider and Patient Assessments in Chemotherapy-Induced Peripheral Neurotoxicity. Neurology 97:e660–e672. 10.1212/WNL.0000000000012300

53. Park SB, Kwok JB, Asher R, et al (2017) Clinical and genetic predictors of paclitaxel neurotoxicity based on patient- versus clinician-reported incidence and severity of neurotoxicity in the ICON7 trial. Annals of Oncology 28:2733–2740. 10.1093/annonc/mdx491

54. Maksten EF, Mørch CD, Jakobsen LH, et al (2025) The course of chemotherapy-induced peripheral neuropathy (CIPN) in hematological patients treated with vincristine, bortezomib, or lenalidomide: the NOVIT study. Support Care Cancer 33:225. 10.1007/s00520-025-09282-3

